# Diversifying deaths: the shifting spectrum of childhood respiratory infectious mortality, 1990–2023: a systematic analysis of the Global Burden of Disease Study 2023

**DOI:** 10.64898/2026.09.01.26361937

**Authors:** Deze Lia, Hao Chen, Yu Miao, Yuxiang Zhang, Xiaotong Wang, Chen Shen

**Affiliations:** Beijing Pediatric Research Institute, Beijing Children’s Hospital, Capital Medical University, Beijing, China

**Keywords:** respiratory infections, child mortality, pathogen spectrum, diversity index, pertussis, tuberculosis, COVID-19, Global Burden of Disease

## Abstract

**Background:** Childhood respiratory infectious deaths are partitioned across four Global Burden of Disease cause modules—26 etiological attributions within lower respiratory infections, tuberculosis, COVID-19, and whooping cough—never jointly reported. Whether the structure of this combined mortality spectrum has changed over time, and with what implications for intervention design, has not been quantified. We assembled and analyzed the integrated spectrum for children and adolescents aged 0– 19 years, 1990–2023.

**Methods:** We integrated Global Burden of Disease Study 2023 (release v8352) estimates into a 29-node spectrum—26 lower respiratory infection etiologies plus tuberculosis, COVID-19, and pertussis— globally and across seven super-regions, with uncertainty propagated by summing bounds. We computed Shannon diversity, Herfindahl concentration, and effective cause counts; phenotyped pandemic-window collapse and rebound per cause; linked pathogen shares to WHO/UNICEF vaccine coverage; and mapped geographic concentration in sub-Saharan Africa and South Asia. Segmented (joinpoint) log-linear models with breakpoints selected by the Bayesian information criterion were fitted to annual totals, Shannon diversity and leading pathogen shares.

**Results:** In 2023 the 29 causes jointly accounted for 965,330 deaths (95% uncertainty interval [UI] 680,096–1,342,437). Shannon diversity rose from 2.336 to 2.711 (+16.1%) between 1990 and 2023; the effective number of causes nearly doubled (5.57 to 9.94), inversely coupled to total deaths (Spearman ρ = −0.997). Whooping cough ranked second (112,954 deaths; 95% UI 64,576–185,708; 11.7%) and showed the spectrum’s only rebound above 100% (−57.4% collapse, +111.0% rebound). Tuberculosis ranked third (87,764; 57,779–124,912; 9.1%) with the highest concentration in sub-Saharan Africa and South Asia (87.1%). COVID-19 entered at rank five (52,899; 47,275–59,183; 5.5%). Nineteen of 29 causes exceeded the poverty-lock threshold (>80.59% of deaths in sub-Saharan Africa plus South Asia). Joinpoint analysis placed breakpoints in total deaths at 2011, 2018 and 2021—the decline slowed from −3.4% to −2.3% per year after 2011, steepened to −7.1% per year in 2018–2021 and rebounded +4.5% per year in 2021–2023—while diversification accelerated to +1.18% per year in 2013–2020; all six leading pathogen shares carried a pandemic-window breakpoint at 2019 and 2021 (S. pneumoniae at 2018 and 2021).

**Conclusions:** Childhood respiratory infectious mortality has become more diverse and more concentrated in poverty as it has declined. Single-pathogen interventions now address a shrinking share; the spectrum’s structure argues for platform interventions—oxygen, antimicrobial access, referral—tailored jointly by age and geography, implying that pathogen-specific strategies alone cannot finish the remaining mortality agenda.

## 1. Introduction

### 1.1 Childhood respiratory infectious deaths are dispersed across four GBD modules

Respiratory infections remain among the leading infectious killers of children and adolescents worldwide. Yet within the Global Burden of Disease (GBD) framework these deaths are partitioned across four cause modules that are never jointly reported: 26 etiological attributions within lower respiratory infections (LRIs), derived through a counterfactual population-attributable-fraction model; tuberculosis; COVID-19; and whooping cough, each estimated as an independent, mutually exclusive cause of death^1 2^. Successive GBD cycles have refined LRI etiological attribution^3 4^, most recently reporting non-COVID-19 LRI mortality for 1990–2021 in 18 etiological categories—including aggregate and residual groupings—with strata for children under 5 years and aged 5–14 years^2^. Two integration precedents exist: the global antimicrobial resistance study spanned infectious syndromes across GBD hierarchical levels^5^, and the GBD 2019 collaboration synthesized the communicable-disease burden of children and adolescents^6^. Neither, however, assembled a single childhood respiratory mortality spectrum at single-pathogen resolution, and neither captured the pandemic window or the post-2021 rebound now observable through 2023.

### 1.2 Three knowledge gaps

First, no study has tracked the diversity of a population-level respiratory pathogen mortality spectrum over time. Ecological diversity metrics have been applied to respiratory pathogens only at other scales—within-species serotype diversity in pneumococcal carriage after conjugate vaccination^7^ and within-host lung-microbiome diversity in critically ill patients with pneumonia^8^—leaving the evenness of the childhood mortality spectrum unquantified. Therefore, we computed Shannon diversity, Herfindahl concentration, and the effective number of pathogens for the full spectrum annually from 1990 to 2023. Second, the COVID-19 pandemic perturbed all respiratory pathogens simultaneously, yet collapse-and-resurgence comparisons have been confined to incidence-based surveillance of viruses^9 10^. Therefore, we used the pandemic window as a natural experiment to phenotype the collapse depth and rebound speed of bacterial, viral, fungal, and pandemic-driven causes jointly on a mortality spectrum. Third, although GBD 2021 incorporated vaccine coverage as an attribution covariate^2^, no explicit pediatric cross-pathogen dose–response geometry linking third-dose pneumococcal and Haemophilus influenzae type b coverage^11^ to spectrum composition has been reported, and the geographic concentration of the residual spectrum has not been formalized as a structural property. Therefore, we mapped each cause’s coverage–mortality gradient and its concentration in sub-Saharan Africa and South Asia.

### 1.3 Contribution statement

**Novelty.** Using GBD 2023, we construct the first integrated 29-node pediatric (0–19 years) respiratory infectious mortality spectrum—26 LRI pathogens plus tuberculosis, COVID-19, and pertussis—for 1990–2023, comprising 965,330 deaths (95% uncertainty interval 680,096–1,342,437) in 2023, and quantify its diversity succession (+16.1% in Shannon H; effective pathogen count 5.57 to 9.94)^12^. **Differentiation.** Relative to GBD 2021’s 18 etiological categories with under-5 and 5–14 year strata^2^, we resolve single pathogens for a dedicated 0–19 population extended through the post-2021 rebound; relative to viral resurgence surveillance^9 10^, our perturbation analysis spans bacteria and viruses on a death spectrum; and relative to prior cross-module integrations^5 6^, we add pathogen-level resolution and the pandemic window. **Implications.** With diversity rising, the vaccine-preventable share shrinking, and 19 of 29 causes concentrated above the poverty-lock threshold, the marginal leverage of single-pathogen interventions declines relative to platform interventions—oxygen therapy, antimicrobial access, and hospital infection control—delivered with age- and geography-specific tailoring.

## 2. Methods

### 2.1 Data source and integration of four cause modules through three estimation interfaces

We conducted a secondary analysis of the Global Burden of Disease Study 2023 (GBD 2023; release v8352), which estimates cause-specific mortality for 204 countries and territories, 1990–2023, using standardized Bayesian hierarchical models ^1 13^. Childhood respiratory infectious mortality is partitioned across four GBD cause modules—26 LRI etiology attributions, tuberculosis, COVID-19 and whooping cough—which are consolidated through three estimation interfaces into a single 29-node spectrum. First, etiologically attributed lower respiratory infection (LRI) deaths were extracted at the risk–effect (rei) level for the 26 single-pathogen causes of GBD 2023 ^1^. Attribution here follows a counterfactual population-attributable fraction (PAF) model ^2 3^; the rei-level interface returns deaths only, so all analyses are mortality-based (metric: Deaths, Number). Second, tuberculosis (cause 297) and whooping cough (cause 339) were extracted from the cause interface, where they are modelled directly (CODEm and specialized ensemble approaches) rather than through PAF attribution ^1 14^. Third, COVID-19 (cause 1048) was extracted for 2020–2023, with zeros for 1990–2019. Because GBD causes of death are mutually exclusive and collectively exhaustive, concatenating the modules introduces no double counting; the two frameworks generate uncertainty differently (PAF-based versus cause-model draws), and module-specific uncertainty intervals (UIs) are treated as structurally heterogeneous.

### 2.2 Definitions, age aggregation, and regional mapping

The primary pediatric total, ages 0–19 years, was defined as the sum of the <5, 5–9, 10–14, and 15–19 year groups; the 20–24 year group was retained solely as a young-adult comparator and never folded into the 0–19 total. UIs were propagated conservatively by summing GBD-reported lower and upper bounds across strata (wider than posterior-draw aggregation), with point estimates summed analogously, keeping totals and shares internally consistent. The extraction grid comprised Global plus the seven GBD super-regions, five age groups, both sexes, and 34 years, with no missing strata. Inputs available only at the 21-region level (the three independently modelled causes) were mapped to super-regions using the standard GBD location hierarchy.

### 2.3 Spectrum-structure metrics

For each location-year we computed, over the 29-node death-share distribution pᵢ, the Shannon diversity index H = −Σ pᵢ ln pᵢ ^12^, the Herfindahl–Hirschman index HHI = Σ pᵢZ, and the effective number of causes effN = 1/HHI. The global time series is reported as effN = 1/HHI and super-region cross-sections as the Shannon number equivalent exp(H) (Hill number, q = 1); every figure and table legend states which metric is used. Decoupling between diversity and burden magnitude was tested by Spearman correlation between annual H and annual total 0–19 deaths (n = 34 years). Because both series trend, we added a first-difference test—Pearson and Spearman correlations between year-to-year changes in diversity (ΔH, and ΔeffN as a concentration-based counterpart) and year-to-year changes in total deaths (n = 33 differences)—to determine whether the level association survives detrending (Table S10). Sensitivity series excluded COVID-19 or tuberculosis (28-node spectra) to check robustness to module composition. Segmented trend (joinpoint) analysis. To localise changes in the pace of mortality decline, diversification and compositional succession, we fitted segmented log-linear (joinpoint) regression models^28^ to the annual global series of total 29-cause deaths, Shannon H, and the death shares of the four leading non-pandemic causes (S. pneumoniae, whooping cough, tuberculosis and K. pneumoniae) and the two established respiratory viruses (influenza and RSV), 1990–2023. For each series, models with 0–3 joinpoints were fitted by exhaustive grid search over admissible joinpoint years—requiring at least three annual observations per segment, including a minimum two-year spacing between adjacent joinpoints— with the number and location of joinpoints selected by the Bayesian information criterion (BIC), the penalty term counting each joinpoint location as an additional estimated parameter (2k + 2 parameters for a model with k joinpoints). We report segment-specific annual percent changes (APC) and the span-weighted average annual percent change (AAPC) for 1990–2023, with 95% confidence intervals from the t distribution with n − (2k + 2) residual degrees of freedom. Models were fitted to central estimates. The complete segment table and reproducible code are provided in Additional file 3.

### 2.4 Pandemic-window perturbation–recovery phenotypes

We treated the COVID-19 period as a natural perturbation experiment on the spectrum, complementing incidence-based observations of asynchronous viral disruption and resurgence ^9 10^. For each node, collapse was the percentage change in global 0–19 deaths from 2019 to 2021 and rebound the change from 2021 to 2023. Nodes were classified a priori into five groups: viruses, bacteria, fungi, other, and pandemic-driven; COVID-19 was assigned singly to the last group, because its zero 2019 baseline renders collapse undefined, and is described by its 2021 peak and subsequent fade. Collapse depth was compared between viruses and bacteria with a two-sided Mann–Whitney U test, and collapse–rebound coupling by Spearman correlation across the 28 nodes with a defined 2019 baseline. All analyses with n < 20 are reported as hypothesis-generating only, in observational wording (“we observed”).

### 2.5 Vaccine dose–response analysis

We linked super-regional change in pathogen share to coverage from WHO/UNICEF Estimates of National Immunization Coverage (WUENIC, 2026 revision), using third-dose pneumococcal conjugate vaccine (PCV3) and third-dose Haemophilus influenzae type b vaccine (Hib3) ^11^. The vaccine-preventable group was redefined as Streptococcus pneumoniae, H. influenzae (type b vaccine targeted), influenza, and pertussis; from 2021, COVID-19 was additionally counted under a second caliber, and both calibers are reported. Because GBD already incorporates coverage as an attribution covariate ^2^, this analysis interrogates the resulting estimates rather than providing independent causal evidence. Coverage responds to introduction with a lag—India introduced PCV nationally only in 2017—and WUENIC zeros denote non-introduction rather than true zero protection; coverage was computed over all countries and over introducing countries only, with population weighting as sensitivity. Super-region medians of 2015 coverage were correlated with the 2010 to 2023 change in the corresponding pathogen’s share (Spearman); with n = 7 super-regions these analyses are descriptive only and subject to ecological fallacy.

### 2.6 Poverty-lock geometry

For each node we computed the share of its 2023 global 0–19 deaths occurring in Sub-Saharan Africa plus South Asia. A node was classified as poverty-locked when this share exceeded 80.59%, the two regions’ combined share of all-29-node 0–19 deaths in 2023; the reference line is recomputed on the 29-node denominator. Sensitivity to dichotomization was examined by also reporting shares continuously. Independence between poverty-lock status and age-tropism quadrant (infant-tropic versus older-child-tropic, by the share-based susceptibility-age index) was tested with a Fisher exact test (n = 14 nodes with computable tropism indices). We state a priori that power at n = 14 is very low; a null result is not interpretable as evidence of independence.

### 2.7 Predefined pathogen groupings

The opportunistic/hospital-associated group comprised Pseudomonas aeruginosa, Staphylococcus aureus, nontuberculous mycobacteria, Acinetobacter baumannii, and Klebsiella pneumoniae—pathogens whose pediatric mortality is plausibly linked to health-care exposure and comorbidity. The vaccine-preventable group was as defined in §2.5. All remaining nodes were analyzed individually. Group shares use the yearly 29-node total as denominator, so changes reflect both numerator dynamics and denominator composition.

### 2.8 Reporting standards, ethics, and software

. The analysis uses publicly available, de-identified, aggregate modelled estimates and was exempt from institutional ethics review. Analyses were performed in Python (pandas, NumPy, SciPy) with two-sided α = 0.05.

## 3. Results

### 3.1 The 29-node spectrum in 2023

In 2023 the 29 mutually exclusive causes jointly accounted for 965,330 (95% uncertainty interval [UI] 680,096–1,342,437) deaths at ages 0–19 years globally, down from 2,573,489 in 1990 (−62.5%). The total combines 711,713 deaths across the 26 lower respiratory infection (LRI) etiologies with three causes modelled elsewhere in the GBD hierarchy: whooping cough (112,954; 95% UI 64,576–185,708), tuberculosis (87,764; 57,779–124,912) and COVID-19 (52,899; 47,275–59,183). *Streptococcus pneumoniae* remained the leading single cause (227,976 deaths; 95% UI 164,927–306,384; 23.6% of the total), but whooping cough ranked second (11.7%) and tuberculosis third (9.1%), ahead of *Klebsiella pneumoniae* (8.8%); COVID-19, a cause that did not exist before 2020, entered at rank 5 (5.5%); influenza and respiratory syncytial virus (RSV), both top-seven causes in 1990, ranked tenth and eleventh. The country-level distribution of the 26 LRI aetiologies in 2023 is provided in Additional file 1 (summarized in Tables S7–S8).

Table 1 ranks all 29 nodes with 2023 deaths, 95% UIs and shares, alongside 1990 shares and rank changes. The spectrum remains overwhelmingly bacterial (23 of 29 nodes), and concentration at the top coexists with a long tail: ranks 16–29 each contribute under 1.7% yet together account for nearly one death in ten. Rank changes over 34 years were modest—the four largest causes in 1990 retained the top four ranks—except for the viral fallers (influenza and RSV, −4 each), *Acinetobacter baumannii* (−4), *Staphylococcus aureus* (+3) and COVID-19 (29 to 5). Relative UI widths are widest for small-count causes such as *Legionella* spp, cautioning against over-reading rank order below approximately rank 15.

**Table 1.**
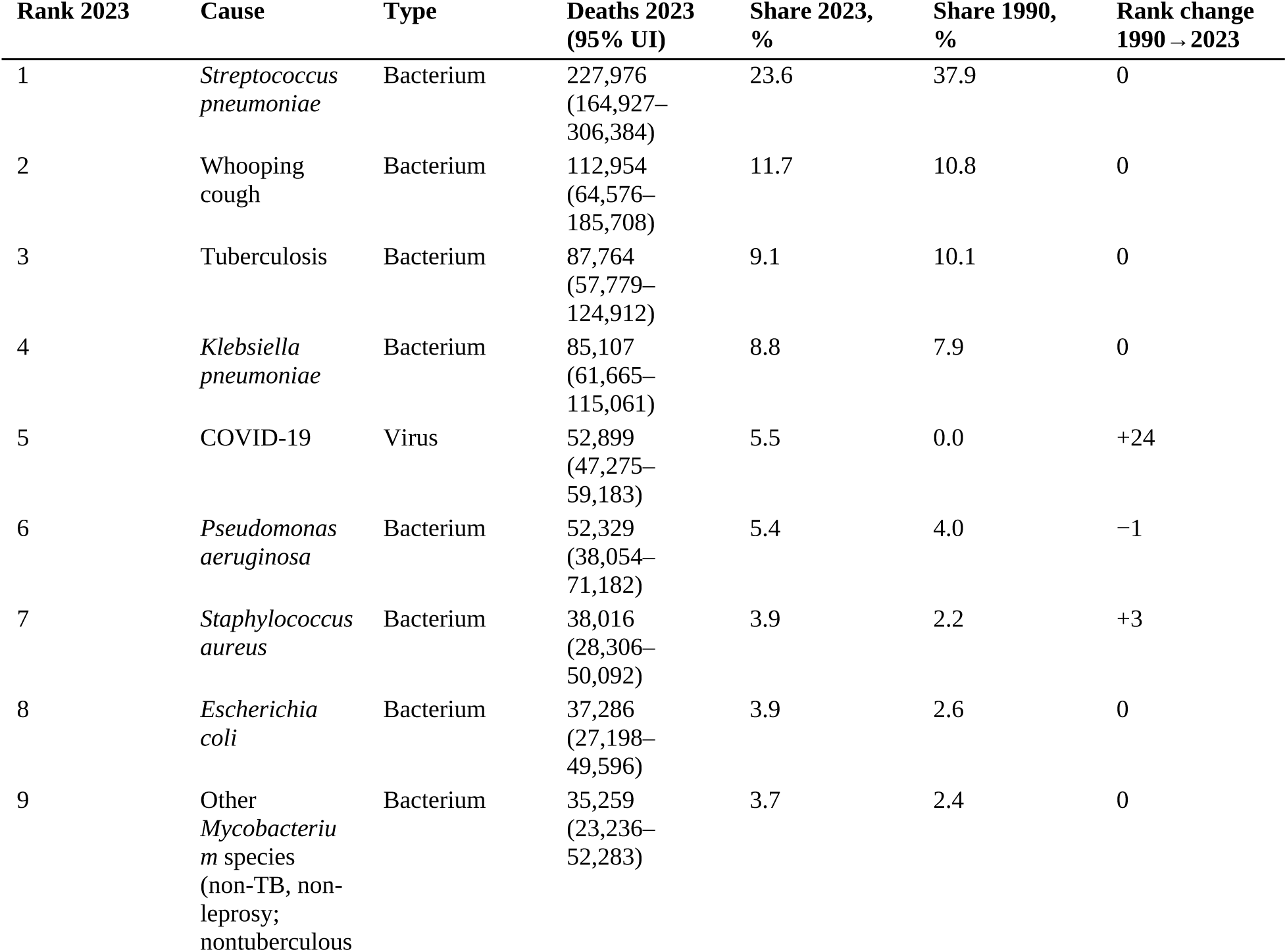
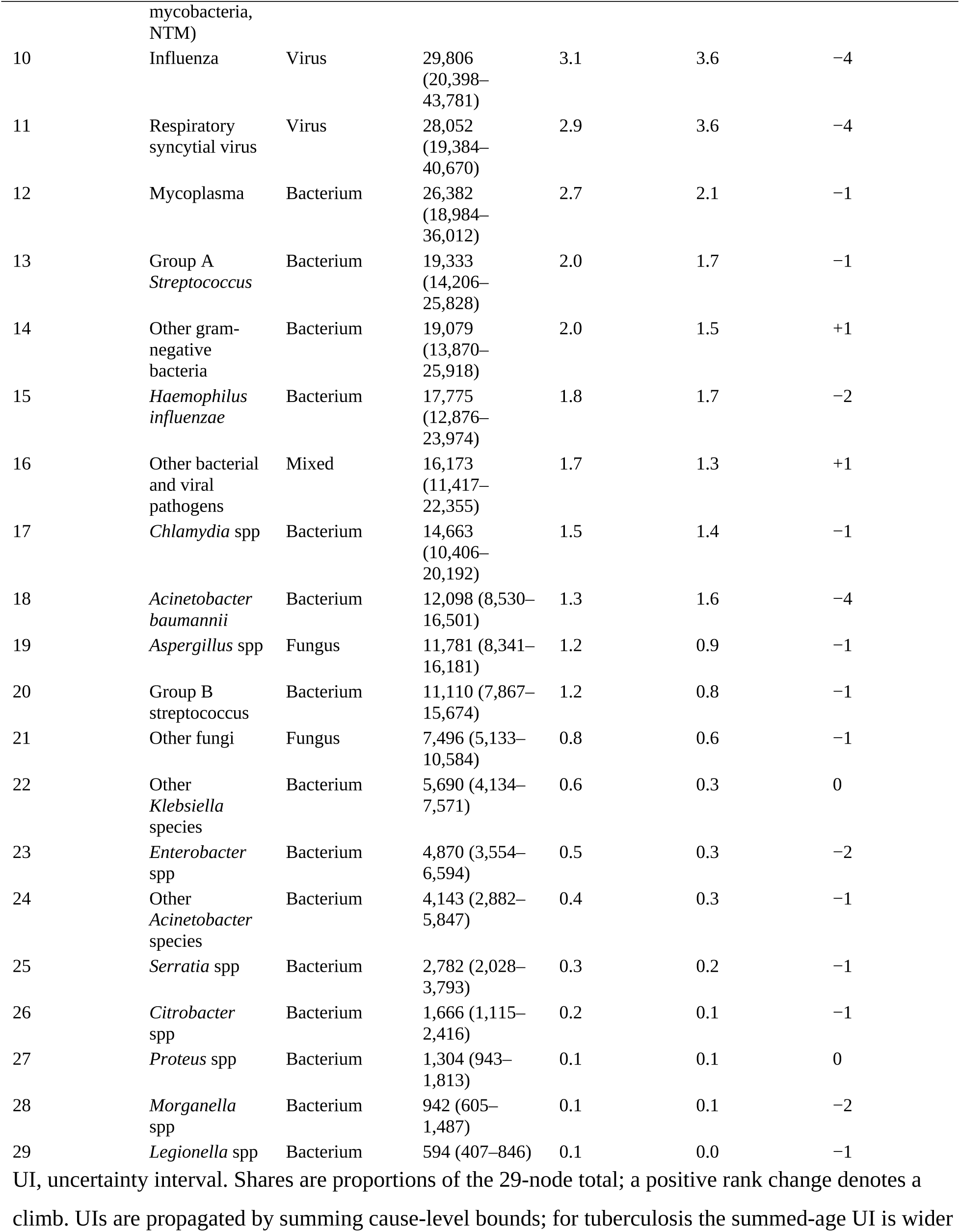
Global deaths from respiratory infectious causes, 29-node spectrum, ages 0–19 years, 2023, with 1990 shares and rank changes Shares below 0.05% are shown as 0.0.

| Rank 2023 | Cause | Type | Deaths 2023 (95% UI) | Share 2023, % | Share 1990, % | Rank change 1990→2023 |
|---|---|---|---|---|---|---|
| 1 | *Streptococcus pneumoniae* | Bacterium | 227,976 (164,927–306,384) | 23.6 | 37.9 | 0 |
| 2 | Whooping cough | Bacterium | 112,954 (64,576–185,708) | 11.7 | 10.8 | 0 |
| 3 | Tuberculosis | Bacterium | 87,764 (57,779–124,912) | 9.1 | 10.1 | 0 |
| 4 | *Klebsiella pneumoniae* | Bacterium | 85,107 (61,665–115,061) | 8.8 | 7.9 | 0 |
| 5 | COVID-19 | Virus | 52,899 (47,275–59,183) | 5.5 | 0.0 | +24 |
| 6 | *Pseudomonas aeruginosa* | Bacterium | 52,329 (38,054–71,182) | 5.4 | 4.0 | −1 |
| 7 | *Staphylococcus aureus* | Bacterium | 38,016 (28,306–50,092) | 3.9 | 2.2 | +3 |
| 8 | *Escherichia coli* | Bacterium | 37,286 (27,198–49,596) | 3.9 | 2.6 | 0 |
| 9 | Other *Mycobacterium* species (non-TB, non-leprosy; nontuberculous mycobacteria, NTM) | Bacterium | 35,259 (23,236–52,283) | 3.7 | 2.4 | 0 |
| 10 | Influenza | Virus | 29,806 (20,398–43,781) | 3.1 | 3.6 | −4 |
| 11 | Respiratory syncytial virus | Virus | 28,052 (19,384–40,670) | 2.9 | 3.6 | −4 |
| 12 | Mycoplasma | Bacterium | 26,382 (18,984–36,012) | 2.7 | 2.1 | −1 |
| 13 | Group A *Streptococcus* | Bacterium | 19,333 (14,206–25,828) | 2.0 | 1.7 | −1 |
| 14 | Other gram-negative bacteria | Bacterium | 19,079 (13,870–25,918) | 2.0 | 1.5 | +1 |
| 15 | *Haemophilus influenzae* | Bacterium | 17,775 (12,876–23,974) | 1.8 | 1.7 | −2 |
| 16 | Other bacterial and viral pathogens | Mixed | 16,173 (11,417–22,355) | 1.7 | 1.3 | +1 |
| 17 | *Chlamydia* spp | Bacterium | 14,663 (10,406–20,192) | 1.5 | 1.4 | −1 |
| 18 | *Acinetobacter baumannii* | Bacterium | 12,098 (8,530–16,501) | 1.3 | 1.6 | −4 |
| 19 | *Aspergillus* spp | Fungus | 11,781 (8,341–16,181) | 1.2 | 0.9 | −1 |
| 20 | Group B streptococcus | Bacterium | 11,110 (7,867–15,674) | 1.2 | 0.8 | −1 |
| 21 | Other fungi | Fungus | 7,496 (5,133–10,584) | 0.8 | 0.6 | −1 |
| 22 | Other *Klebsiella* species | Bacterium | 5,690 (4,134–7,571) | 0.6 | 0.3 | 0 |
| 23 | *Enterobacter* spp | Bacterium | 4,870 (3,554–6,594) | 0.5 | 0.3 | −2 |
| 24 | Other *Acinetobacter* species | Bacterium | 4,143 (2,882–5,847) | 0.4 | 0.3 | −1 |
| 25 | *Serratia* spp | Bacterium | 2,782 (2,028–3,793) | 0.3 | 0.2 | −1 |
| 26 | *Citrobacter* spp | Bacterium | 1,666 (1,115–2,416) | 0.2 | 0.1 | −1 |
| 27 | *Proteus* spp | Bacterium | 1,304 (943–1,813) | 0.1 | 0.1 | 0 |
| 28 | *Morganella* spp | Bacterium | 942 (605–1,487) | 0.1 | 0.1 | −2 |
| 29 | *Legionella* spp | Bacterium | 594 (407–846) | 0.1 | 0.0 | −1 |

UI, uncertainty interval. Shares are proportions of the 29-node total; a positive rank change denotes a climb. UIs are propagated by summing cause-level bounds; for tuberculosis the summed-age UI is wider [illegible]

### 3.2 Diversity rose while deaths fell

Herfindahl–Hirschman index (HHI) fell from 0.179 to 0.101

Figure 1 displays the decoupling: total deaths trend downward (panel a) while Shannon H rises (panel b), with HHI confirming that de-concentration is not index-specific (panel c). Dashed traces show the two 28-node sensitivity series, which bracket the main series without altering its slope or pandemic-window behavior. The shaded pandemic years do depart briefly from the monotonic decline: after the 2021 trough (893,387 deaths), total deaths and diversity rose together for two consecutive years (928,035 in 2022; 965,330 in 2023), yet the 34-year inverse correlation is essentially unaffected (ρ = −0.997). The super-region panel adds a gradient consistent with an epidemiological transition: where mortality has fallen furthest the spectrum is most even. Rising diversity alongside falling deaths is therefore structural over the full series, not an artefact of the added causes or the pandemic window.

**Figure 1.**
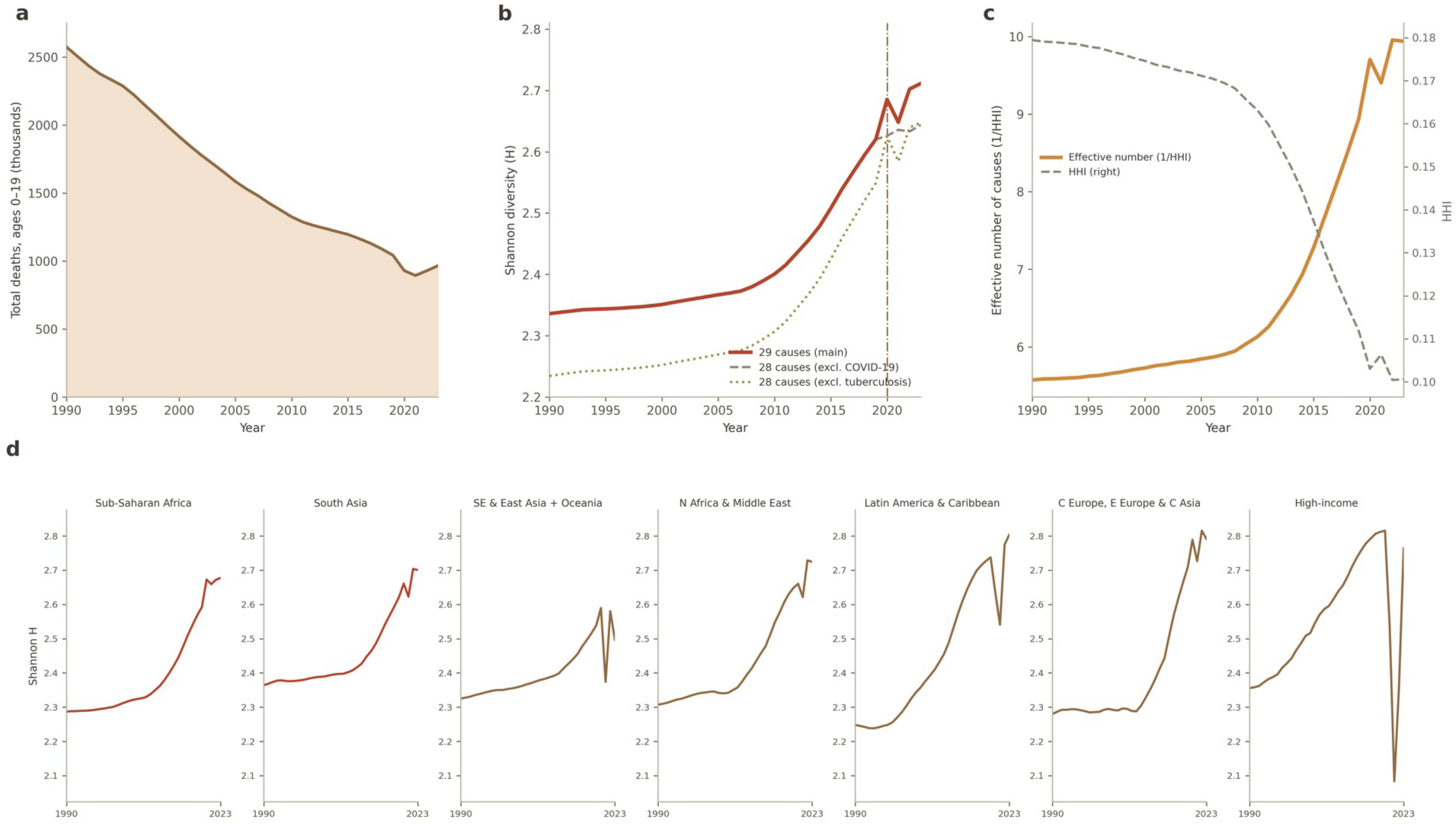
Diversity–mortality decoupling among children aged 0–19 years, 1990–2023. (a) Total deaths summed over the 29 causes; (b) Shannon diversity H of the 29-cause death-share distribution; dashed traces are the 28-cause sensitivity series excluding COVID-19 or tuberculosis, and the vertical line marks 2020; (c) effective number of causes (1/HHI), with the Herfindahl–Hirschman index (HHI) on the secondary axis; (d) Shannon H by GBD super-region, 1990–2023.

Joinpoint analysis resolved these aggregate movements into phases (Additional file 3). Total 29-cause deaths declined by −3.4% per year (95% CI −3.5 to −3.3) in 1990–2011, with the pace of decline slowing to −2.3% per year (−2.6 to −1.9) in 2011–2018, steepening to −7.1% per year (−8.2 to −6.0) across the pandemic onset in 2018–2021, and rebounding +4.5% per year (2.3 to 6.8) in 2021–2023 (AAPC 1990–2023 −3.0% per year, −3.1 to −2.9). Shannon diversity rose by +0.09% per year (0.07–0.12) in 1990–2008, accelerating to +0.65% (0.55–0.74) in 2008–2013 and +1.18% (1.09–1.26) in 2013–2020, before decelerating to +0.61% (0.37–0.85) in 2020–2023 (AAPC +0.45% per year, 0.43–0.47). Diversification thus accelerated through the vaccine scale-up era and into the pandemic window while mortality decline first slowed and then steepened, reinforcing at annual resolution that the two movements are structurally decoupled.

### 3.3 Diversification is near-universal at the national level

To test whether diversification is an aggregation artefact driven by a few large countries, we computed national Shannon diversity over the 26 LRI aetiologies for each of the 204 countries and territories at the five time points of Additional file 1 (1990, 2010, 2019, 2021 and 2023; Table S9; 2023 values are mapped in Figure 4c). National H rose between 1990 and 2023 in 203 of 204 countries (99.5%); the sole exception was Somalia (ΔH = −0.006). The cross-country median H rose from 2.11 in 1990 to 2.62 in 2023 (2.30 in 2010; 2.59 in 2019; 2.57 in 2021). Diversification was faster where baseline diversity was lowest (Spearman correlation between 1990 H and ΔH, ρ = −0.19, p = 0.007), yet cross-country dispersion widened (interquartile range of H, 0.132 in 1990 versus 0.259 in 2023)—convergence in slope, divergence in level. The pandemic window showed a scale contrast: national H dipped between 2019 and 2021 in 70.6% of countries and recovered by 2023 in 86.3%, even though global H did not dip (§3.2). Finally, the cross-country correlation between ΔH and the 1990 to 2023 change in log deaths was weak and non-significant (ρ = −0.12, p = 0.077): how much a country’s mortality fell explains almost none of how much its spectrum diversified.

### 3.4 Compositional succession: prevention versus opportunity

The vaccine-preventable group (*S. pneumoniae*, *H. influenzae*, influenza and whooping cough) fell from 54.0% of all deaths in 1990 to 40.2% in 2023 (−13.8 percentage points [pp]); under the alternative definition including COVID-19 from 2021 onwards, the 2023 share was 45.7% (Figure 2). The opportunistic and hospital-associated group (*P. aeruginosa*, *S. aureus*, nontuberculous mycobacteria, *A. baumannii*, *K. pneumoniae*) rose from 18.1% to 23.1% (+5.0 pp). *S. pneumoniae* declined from 37.9% to 23.6% (−14.3 pp) yet retained rank 1; tuberculosis (10.1% to 9.1%) and whooping cough (10.8% to 11.7%) held ranks 3 and 2 with near-stable shares. Because shares are relative, rightward movement denotes redistribution within a shrinking total, not resurgence.

**Figure 2.**
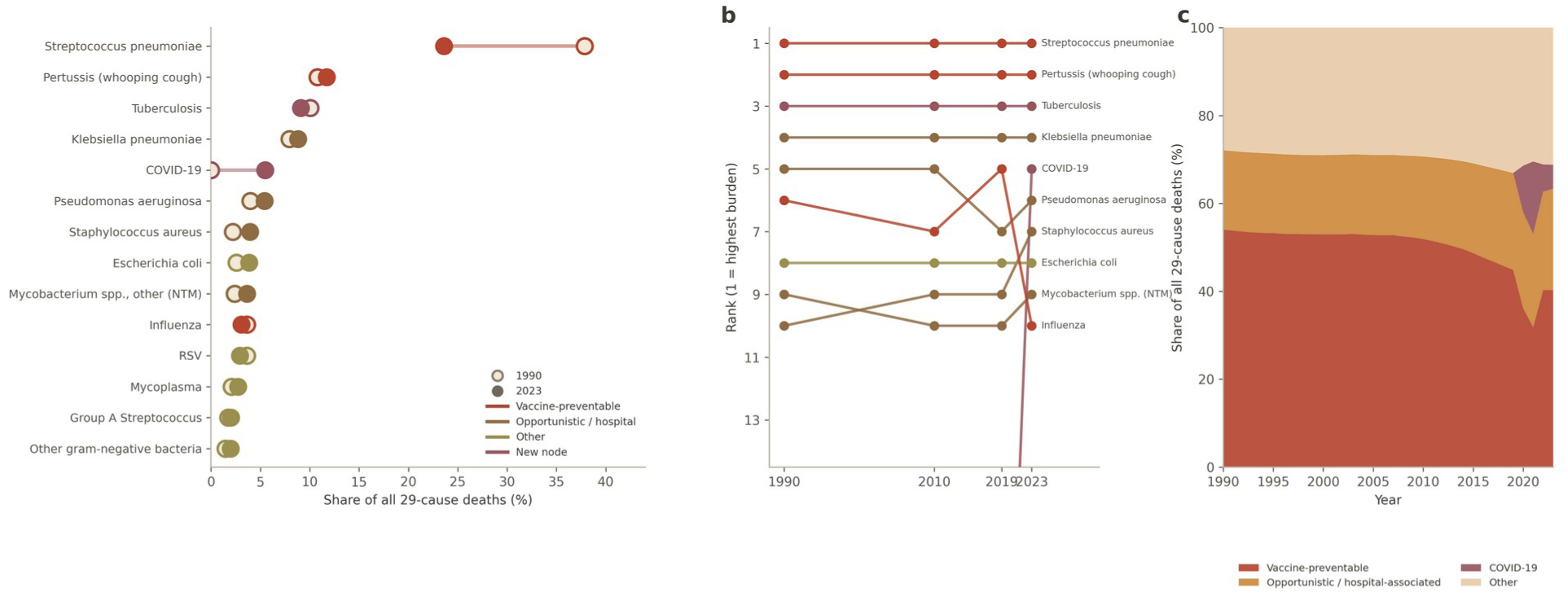
Compositional succession of the 29-cause spectrum. (a) Share of all 29-cause deaths in 1990 versus 2023 for the 14 highest-burden causes (dumbbells), coloured by group (vaccine-preventable, opportunistic or hospital-associated, other, and new nodes); (b) rank trajectories of the ten highest-burden causes at 1990, 2010, 2019 and 2023; (c) annual composition by channel (vaccine-preventable; opportunistic or hospital-associated; COVID-19; other), stacked percentage shares, 1990–2023.

Figure 2 plots each cause’s 1990 and 2023 shares as dumbbells for the 14 highest-burden nodes. The dominant feature is the long leftward arc of *S. pneumoniae*, driving most of the vaccine-preventable contraction, above consistently rightward opportunistic arcs; the gap between the two group totals narrowed from 35.9 pp to 17.2 pp. The three added causes occupy distinct positions: whooping cough and tuberculosis sit in the upper, slowly contracting band, whereas COVID-19 appears as a de novo 2023-only point. The two-calibre vaccine-group total quantifies how much residual vaccine-preventable burden in 2023 reflects the newly endemic coronavirus rather than the four classic targets.

Segmented trends add temporal resolution to this succession (Additional file 3). The S. pneumoniae share declined −0.3% per year (95% CI −0.4 to −0.3) in 1990–2011, then −3.7% per year (−3.8 to −3.5) in 2011–2018 and −7.0% per year (−7.5 to −6.4) in 2018–2021, before rebounding +3.6% per year (2.5 to 4.8) in 2021–2023: the erosion of the dominant pathogen was concentrated in 2011–2021, followed by partial share recovery. The whooping cough share drifted upward +0.5% per year (0.2–0.8) through 1990–2019, collapsed −25.8% per year (−30.6 to −20.6) in 2019–2021 and rebounded +44.4% per year (31.0–59.1) in 2021–2023, the steepest V-shape on the spectrum. The tuberculosis share rose +1.2% per year (1.0–1.4) in 1990–1999, drifted downward −0.7% per year (−0.8 to −0.7) in 1999–2019, jumped +5.5% per year (4.3–6.8) in 2019–2021 and fell back −8.5% per year (−10.0 to −6.9) in 2021–2023 (AAPC −0.3% per year, −0.4 to −0.3); the K. pneumoniae share edged down −0.2% per year (−0.2 to −0.1) in 1990–2009, rose +1.1% per year (1.1–1.2) in 2009–2019, dipped −3.0% per year (−3.4 to −2.6) in 2019–2021 and recovered +4.5% per year (4.0–5.1) in 2021–2023 (AAPC +0.3% per year, 0.3 to 0.4). The influenza and RSV shares rose in 2011–2019 and 2009–2019 (+4.0% and +3.1% per year), collapsed by −33.4% and −35.7% per year in 2019–2021, and rebounded +20.6% and +22.1% per year in 2021–2023 without regaining their 2019 levels by 2023 (influenza 5.3% in 2019 versus 3.1% in 2023; RSV 5.2% versus 2.9%). All six leading shares carry a pandemic-window breakpoint at 2019 and 2021 (S. pneumoniae at 2018 and 2021), marking the pandemic window as the dominant structural break in the spectrum’s composition.

### 3.5 Perturbation–recovery phenotypes during the COVID-19 window

Median 2019 to 2021 collapse was −63.9% for the two established viruses (influenza, RSV) versus −17.1% for the 23 bacteria (Mann–Whitney U p = 0.0067, hypothesis-generating: with two viruses against 23 bacteria, the two-sided p value is bounded below at 0.007; Figure 3, Table 2). Across the 28 nodes with a defined 2019 baseline, we observed that deeper collapse coincided with faster relative rebound (Spearman ρ = −0.574, p = 0.0014), yet both viruses remained well below baseline in 2023 (net −46.1% influenza, −48.5% RSV), consistent with the multi-virus collapse and asynchronous, sometimes off-season, viral resurgence documented in post-pandemic incidence surveillance^9 10 15 16^. Tuberculosis was the most inertial cause: it collapsed least (−4.4%) and did not rebound at all, declining a further 7.9% from 2021 to 2023 (net −12.0%; rebound rank 28 of 28). Whooping cough showed the deepest bacterial V-shape, collapsing 57.4% to a 2021 trough of 53,538 deaths and rebounding 111.0% to 112,954 by 2023 (net −10.1%)—the only node whose rebound exceeded 100%, ranking first on both relative and absolute rebound. This mortality rebound parallels incidence surveillance: US pertussis cases in 2024 were five- to six-fold those of 2023^17^, the EU/EEA notification rate reached 54.9 per 100,000 in 2024, roughly eight times the 2023 rate^18^, and a marked resurgence was documented in France^19^. *Mycoplasma pneumoniae* (GBD etiology label “Mycoplasma”) occupied coordinates (−17.1%, +16.1%), almost exactly the median bacterial phenotype.

**Figure 3.**
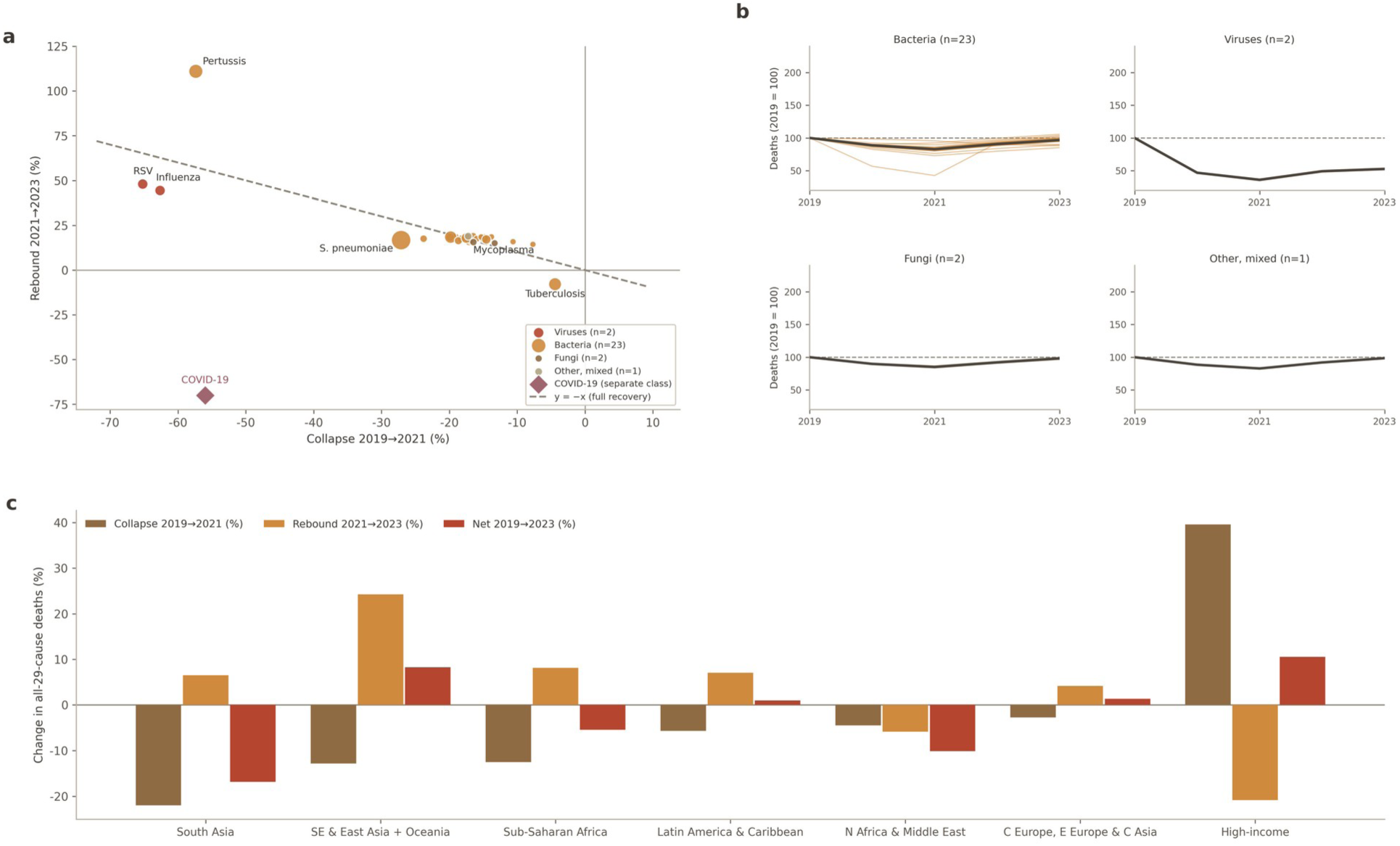
Pandemic-window perturbation and recovery across the 29 causes. (a) Percentage change in deaths among children aged 0–19 years from 2019 to 2021 (x-axis) against the change from 2021 to 2023 (y-axis); points are coloured by cause group and sized by 2019 deaths; the dashed diagonal marks full recovery (y = −x); COVID-19 is plotted separately on a peak-versus-fade basis because its 2019 baseline is zero; (b) deaths indexed to 2019 (= 100), 2019–2023, faceted by cause group, with the group median overlaid; (c) super-regional decomposition of the 2019 to 2021 collapse, the 2021 to 2023 rebound, and the net 2019 to 2023 change in all-29-cause deaths.

**Table 2.** Pandemic-window perturbation–recovery phenotypes, representative causes, global, ages 0–19 years.

| **Cause** | **Type** | **Collapse 2019→2021, %** | **Rebound 2021→2023, %** | **Net 2019→2023, %** | **Rebound rank (of 28)** |
|---|---|---|---|---|---|
| Respiratory syncytial virus | Virus | −65.2 | +48.0 | −48.5 | 2 |
| Influenza | Virus | −62.6 | +44.4 | −46.1 | 3 |
| Whooping cough | Bacterium | −57.4 | +111.0 | −10.1 | 1 |
| *Streptococcus pneumoniae* | Bacterium | −27.1 | +16.7 | −14.9 | 20 |
| *Acinetobacter baumannii* | Bacterium | −23.8 | +17.5 | −10.4 | 15 |
| *Haemophilus influenzae* | Bacterium | −19.9 | +18.3 | −5.2 | 12 |
| *Klebsiella pneumoniae* | Bacterium | −19.8 | +18.4 | −5.0 | 10 |
| *Chlamydia* spp | Bacterium | −19.1 | +18.4 | −4.2 | 11 |
| *Pseudomonas aeruginosa* | Bacterium | −17.5 | +17.9 | −2.7 | 13 |
| Mycoplasma | Bacterium | −17.1 | +16.1 | −3.7 | 23 |
| *Aspergillus* spp | Fungus | −16.5 | +15.6 | −3.4 | 25 |
| *Staphylococcus aureus* | Bacterium | −14.6 | +17.1 | +0.1 | 18 |
| *Legionella* spp | Bacterium | −7.7 | +14.4 | +5.6 | 27 |
| Tuberculosis | Bacterium | −4.4 | −7.9 | −12.0 | 28 |
| COVID-19 | Pandemic-driven | Not applicable | Not applicable | Not applicable | — |

COVID-19 is reported separately because its 2019 baseline is zero and collapse is undefined: deaths rose from 99,113 in 2020 to a peak of 147,942 in 2021 (16.6% of the 29-node total that year), then receded to 52,899 by 2023 (−64.2% from peak), a trajectory consistent with transition towards endemicity^20^. Figure 3 plots collapse against rebound for all 29 nodes, with points sized by 2019 deaths and colored by cause group; COVID-19 appears on a separate peak-versus-fade axis. The negative diagonal cloud visualises the collapse–rebound coupling, with whooping cough isolated in the deep-collapse/fast-rebound quadrant and tuberculosis alone in the no-recovery corner; point size separates speed from magnitude—*M. pneumoniae*’s modest percentage rebound (+3,657 deaths) ranked tenth in absolute terms.

Causes are ordered by collapse depth; COVID-19 is listed separately because its 2019 baseline is zero (2021 peak 147,942 deaths; −64.2% from peak to 2023). Rebound rank is by relative (percentage) rebound among the 28 nodes with defined baselines; absolute-increment ranks differ for high-burden causes. Percentage changes are computed from unrounded values.

Table 2 quantifies phenotypes for 14 representative causes plus COVID-19, adding net 2019 to 2023 change to distinguish recovery speed from completeness. Fast viral rebounds still left deaths nearly half below baseline, whereas several bacteria (*S. aureus*, *Legionella* spp) ended at or above 2019 levels. Relative and absolute rebound ranks diverge for high-burden causes—Mycoplasma ranks 23rd relatively but tenth absolutely—so both metrics are reported. The complete 29-node matrix appears in Supplementary Table S2.

These perturbations far exceeded natural year-to-year fluctuation. Benchmarked against 2017–2019 natural-variability bands, the pandemic-window deviations of all 26 LRI aetiologies fell outside their bands in both 2020 and 2021, and 23 of 26 remained outside in 2023 (Additional file 2, Supplementary Tables S11–S13 and Supplementary Figures S1–S2). M. pneumoniae was a mid-depth case: on the V-shape ranking that combines trough depth with the rebound to 2023 it placed 19th of 26 aetiologies, and its 2021 deviation from the 2017–2019 mean (−18.9%) was far shallower than respiratory syncytial virus (−65.2%) or influenza (−62.1%).

### 3.6 Vaccine coverage and the pneumococcal share: a null dose–response

The pneumococcal share of the 29-node total fell between 2010 and 2023 (35.3% to 23.6%), alongside contraction of the broader vaccine-preventable group (§3.4), as global third-dose pneumococcal conjugate vaccine (PCV3) coverage rose^11^. The super-region dose–response was nonetheless null: we observed no rank correlation between 2015 median PCV3 coverage and the 2010 to 2023 change in pneumococcal share (Spearman ρ = +0.108, p = 0.818, n = 7, 29-node denominator; hypothesis-generating). We report this negative finding without qualification. With seven observational units, heterogeneous introduction dates and no within-region variation, the analysis is descriptive and cannot separate vaccine effects from secular decline; it is subject to ecological fallacy, and a WUENIC value of 0 denotes non-introduction rather than true zero coverage^11^.

### 3.7 Poverty lock-in of the residual spectrum

Sub-Saharan Africa and South Asia jointly accounted for 80.59% of all 29-node deaths in 2023; causes above this reference share were classified as poverty-locked (Figure 4). Nineteen of 29 nodes exceeded the line, most extremely tuberculosis (87.1%), *Chlamydia* spp (86.0%) and *A. baumannii* (84.3%); whooping cough (82.7%) and *S. pneumoniae* (81.7%) were also locked. COVID-19 (54.0%) lay well below the reference and was not locked, and *Legionella* spp (29.8%) was the only cause concentrated outside the two regions. Because the reference share fell from 81.44% (26-pathogen denominator) to 80.59% (29-node denominator), the five causes whose shares lie between the two lines—influenza (81.1%), *Escherichia coli* (80.9%), *Enterobacter* spp (80.9%), *Serratia* spp (80.7%) and *H. influenzae* (80.6%)—newly cross it, whereas *S. pneumoniae* (81.7%) already exceeded the former 81.44% line; the classification is denominator-relative.

**Figure 4.**
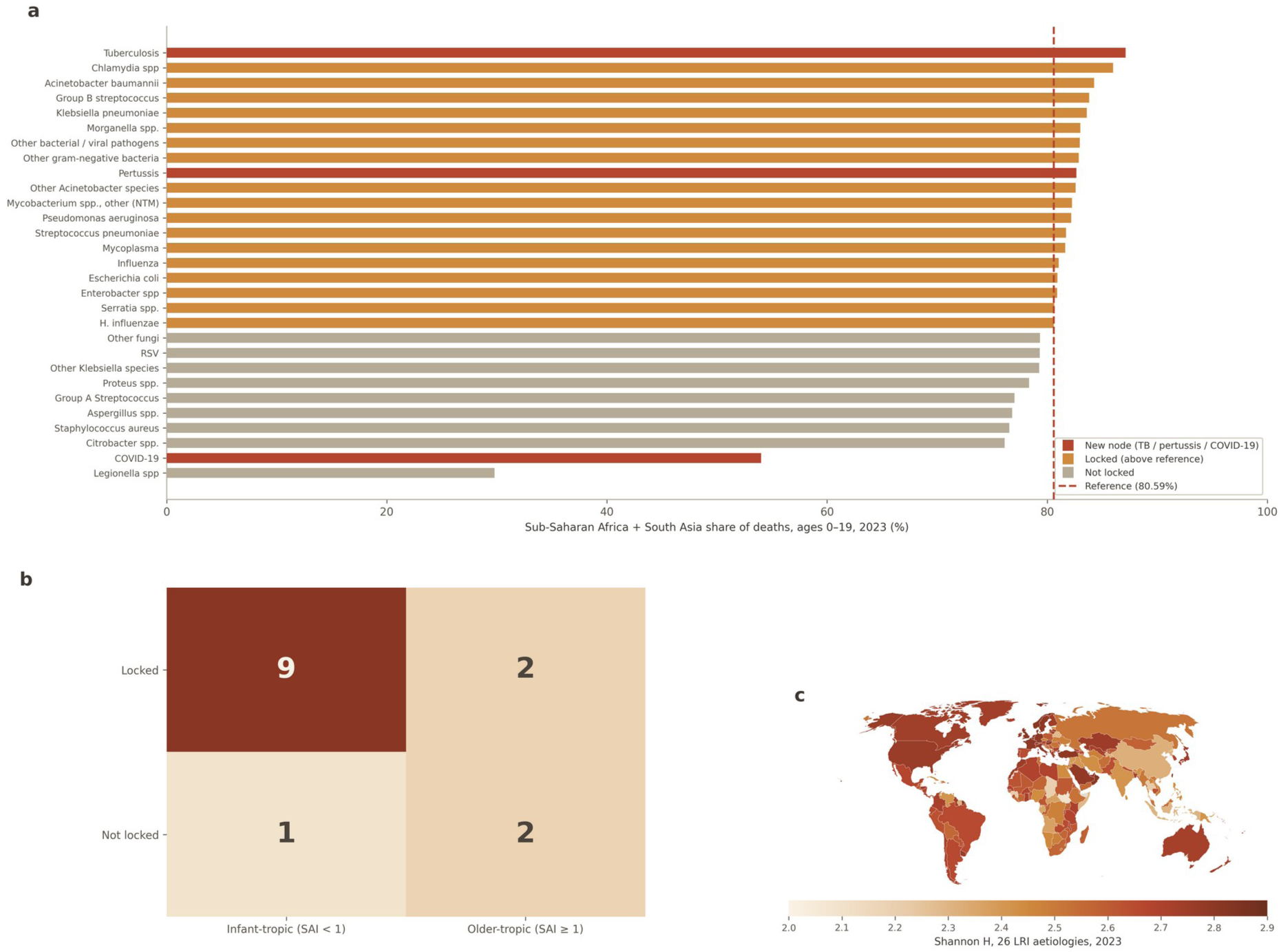
Poverty lock-in and its correlates. (a) Sub-Saharan Africa plus South Asia share of each cause’s deaths among children aged 0–19 years, 2023, ranked, against the 80.59% reference line (dashed); causes above the line are classified as poverty-locked; the three added nodes (tuberculosis, pertussis, and COVID-19) are highlighted; (b) cross-tabulation of poverty lock-in by age tropism for the 14 causes with computable School-Age Index (SAI) values (infant-tropic, SAI < 1; older-tropic, SAI ≥ 1); cell values are counts of causes (Fisher exact p = 0.176); (c) national Shannon diversity of the 26 LRI aetiologies among children aged 0–19 years, 2023; grey indicates no estimate.

Figure 4 ranks all 29 causes by their sub-Saharan Africa plus South Asia share against the 80.59% reference line, with locked causes highlighted. The gradient is continuous rather than bimodal, and the locked set is taxonomically heterogeneous—spanning neonatal bacteria, hospital-associated Gram-negatives, the classic vaccine targets and the added endemic causes—suggesting that geographic concentration reflects health-system context rather than pathogen biology. Tuberculosis, the most inertial cause in the perturbation analysis (§3.5), is also the most strongly locked: the cause least disturbed by the pandemic is the one most confined to the highest-poverty regions, whereas COVID-19 shows the diffuse footprint expected of a recently pandemic virus. Across the 14 causes with computable age-tropism indices, we observed no detectable association between poverty lock-in and age tropism (Fisher exact p = 0.176; hypothesis-generating, as power at n = 14 is minimal; §2.6, §4.2).

## 4. Discussion

### 4.1 Principal findings

By consolidating four GBD 2023 cause modules through three estimation interfaces into a single 29-node spectrum, this analysis shows that childhood respiratory infectious mortality has diversified as it has declined. Four findings stand out. First, the 2023 spectrum comprises 965,330 deaths (95% UI 680,096– 1,342,437) at ages 0–19 years, with whooping cough (11.7%) and tuberculosis (9.1%) ranking second and third—burdens invisible to analyses confined to LRI etiologies—and COVID-19 entering at rank 5 (5.5%). Second, diversity rose while deaths fell: Shannon H increased 16.1% (2.336 to 2.711), the effective number of causes nearly doubled (5.57 to 9.94), and diversity and total burden were near-perfectly inversely coupled (ρ = −0.997). Third, the pandemic window separated causes into distinct perturbation–recovery phenotypes: whooping cough showed the deepest bacterial V-shape (−57.4% collapse; +111.0% rebound, the only rebound above 100%), while tuberculosis was wholly inertial (−4.4%; −7.9%). Fourth, the residual spectrum is poverty-locked: 19 of 29 causes derived more than 80.59% of deaths from sub-Saharan Africa and South Asia, most extremely tuberculosis (87.1%), whereas COVID-19 (54.0%) was not locked.

The national panel adds a fifth, qualifying finding. Diversification is not an aggregation artefact of a few large countries: national Shannon diversity rose in 203 of 204 countries between 1990 and 2023, across all baseline diversity levels, burdens and regions (§3.3). The same panel disciplines the headline decoupling statistic: the near-perfect inverse correlation between annual diversity and annual deaths (ρ = −0.997) is a correlation between two trending time series and is therefore vulnerable to spurious-trend criticism, and the appropriate cross-sectional check does not reproduce it—across countries, the 1990 to 2023 change in log deaths explained almost none of the variation in diversification (ρ = −0.12, p = 0.077). A first-difference check on the global series points the same way: the near-perfect inverse level correlation does not survive detrending—across the 33 year-to-year differences it collapses and flips sign (ΔH versus Δdeaths: Pearson r = +0.22, p = 0.216; Spearman ρ = +0.49, p = 0.004), and the 26-node and effective-number counterparts behave the same (Table S10)—confirming that the level statistic is carried by the shared secular trend rather than by any year-to-year inverse coupling. We therefore interpret diversification not as a mechanical consequence of each country’s own pace of mortality decline but as a near-universal accompaniment of the childhood epidemiological transition. This qualification strengthens rather than weakens the policy reading: because diversification is ubiquitous rather than confined to fast-declining settings, the structural case for platform interventions (§4.2) applies to virtually every national context, while the rate at which a country’s spectrum diversifies must be understood as driven by factors other than its aggregate mortality trend.

### 4.2 Policy implications of diversity succession

The near-doubling of the effective number of causes has direct resource implications. When deaths concentrate on a few dominant pathogens, single-pathogen vaccines address a large fraction of mortality; as the spectrum evened out, the marginal share addressable by any one such intervention shrank. The contraction of the vaccine-preventable group (54.0% to 40.2%; 45.7% including COVID-19) is therefore a structural transition rather than vaccine failure—the compositional signature of prevention success— and it shifts the marginal cost-effectiveness balance towards platform interventions. In sub-Saharan Africa and South Asia, where four-fifths of deaths now occur, oxygen systems with pulse oximetry triage, antimicrobial access, and referral of severe cases act on the shared final pathway of many causes simultaneously, plausibly at lower cost per death averted than introducing a further pathogen-specific vaccine against a cause contributing a single-digit share. We observed no detectable association between poverty lock-in and age tropism (Fisher exact p = 0.176), although with 14 evaluable causes this test had minimal power and the null should not be read as evidence of independence. Which children die where and at what age are therefore treated as two separate axes: interventions should be specified on a two-dimensional matrix of age tropism by geographic concentration rather than deployed uniformly. The rising opportunistic share (18.1% to 23.1%) is likewise denominator restructuring within a shrinking total, not an emerging epidemic of hospital pathogens; *Legionella* spp was the sole pre-existing cause (other than COVID-19, which did not exist before 2020) with more deaths in 2023 than in 1990, rising from 0.01% to 0.1% of the spectrum (594 deaths, 95% UI 407–846). This is a small-base increase compatible with improved diagnosis and environmental or water-system exposure rather than a true surge.

### 4.3 Mechanisms underlying perturbation phenotypes

Within the bacterial group, collapse depth stratified plausibly by transmission biology. *M. pneumoniae* collapsed 17.1%—exactly the bacterial median—consistent with prolonged carriage buffering it against interrupted transmission, in line with its delayed global re-emergence in surveys ^21 22^. *S. pneumoniae* collapsed more deeply (−27.1%), where reduced carriage likely compounded the ongoing PCV-driven decline. Tuberculosis was the most inertial cause (−4.4% collapse; no rebound), as expected for a chronic infection whose annual deaths reflect accumulated incidence over years rather than contemporaneous transmission. Benchmarked against 2017–2019 natural-variability bands (Additional file 2, Supplementary Table S11), every LRI aetiology breached its band in 2020–2021, yet breach depth tracked the same transmission gradient—from −65.2% (respiratory syncytial virus) and −62.1% (influenza) through −33.6% (S. pneumoniae) and −18.9% (M. pneumoniae) to −4.4% for tuberculosis— consistent with suppressed contemporaneous transmission rather than estimation artefact. Whooping cough combined the deepest bacterial V-shape (−57.4% to +111.0%) with a well-documented real-world counterpart: disrupted diphtheria–tetanus–pertussis vaccination and reduced circulation during the pandemic were followed by the multi-country resurgence in incidence shown in Section 3.5 ^19 17 18^. COVID-19, by contrast, peaked at 147,942 deaths in 2021 and receded to 52,899 by 2023, a trajectory consistent with transition towards endemicity ^20^. Its modest standing within the pediatric spectrum accords with population-based evidence that confirmed SARS-CoV-2 deaths in children are rare—25 among 12,023,568 under-18s in England during the first pandemic year ^23^.

### 4.4 Vaccine-era geometry and the null dose–response

The null super-region dose–response (29-node denominator: PCV3 ρ = +0.108, p = 0.818; Hib3 ρ = −0.036, p = 0.939; n = 7) should be read positively. The pneumococcal share fell in all seven super-regions while global PCV3 coverage rose ^11^, and conjugate-vaccine eras are known to restructure pneumococcal populations through serotype replacement alongside herd immunity ^24 25^, a within-species precedent for the spectrum-level transition we observe. Seven observational units with heterogeneous introduction dates cannot support rank correlation, and coverage already enters the GBD attribution model as a covariate ^2^. National-level interrupted time series around documented introduction cohorts— for example India’s 2017 PCV rollout—offer a more powered design to test the share-level response, and such analyses are a priority for future work.

### 4.5 Methodological placement of tuberculosis and pertussis

Including tuberculosis and pertussis in a respiratory mortality spectrum is justified structurally and clinically. Tuberculosis and LRI sit at the same level of the GBD cause hierarchy under a common communicable-disease parent ^1^; pertussis, although classified under other infectious diseases, is clinically a respiratory, vaccine-preventable disease. Cross-module integration follows precedents that assembled infection syndromes across GBD levels ^5 6^. Unlike the 26 LRI etiologies, tuberculosis is modelled directly rather than through PAF attribution, a framework difference we declare rather than reconcile. Comparisons with external estimates must respect caliber: WHO’s Global Tuberculosis Report 2024 estimates 191,000 deaths among under-15s (166,000 HIV-negative; 25,000 HIV-positive) ^26^, against our 87,764 at ages 0–19—differences reflecting estimation framework and HIV stratification more than age bands. Similarly, Yeung and colleagues estimated 160,700 pertussis deaths among under-5s in 2014 ^27^; our 2023 estimate of 112,954 at ages 0–19 spans a wider age band a decade later and is consistent with continued decline.

### 4.6 Limitations

Several limitations qualify these findings. First, the rei-level interface provides deaths only; incidence and lethality effects cannot be separated. Second, etiology-level estimates are publicly released nationally only for the most recent year; this paper accompanies a 204-country dataset at five time points (1990, 2010, 2019, 2021, 2023; Additional file 1), but the full annual 1990–2023 series remains available only at the super-region level (n = 7), limiting longitudinal and dose–response power below the super-region. Third, LRI pathogen shares derive from a modelled counterfactual PAF framework ^2^, whose mutual-exclusivity assumption abstracts from clinical co-infection: real pneumonias are frequently polymicrobial ^8^, so the spectrum partitions deaths that clinically co-occur and its structure would differ under a multi-cause attribution. Fourth, uncertainty intervals are structurally heterogeneous across modules (PAF-based versus cause-model draws) and were propagated by summing bounds, overstating true uncertainty of totals. Fifth, correlations at n = 7 super-regions and n = 14 tropism-indexed causes are hypothesis-generating only. Sixth, WUENIC zeros denote non-introduction, not zero coverage ^11^. Seventh, aggregate correlations cannot establish individual-level causation. Eighth, cross-round comparisons are complicated by differing granularity between GBD 2021 (18 categories) ^2^ and GBD 2023. Ninth, COVID-19 contributes only four observation years, so its phenotype and endemic trajectory remain provisional. Finally, 2023 pertussis estimates likely precede the 2024 resurgence ^19 17 18^ and may be revised upward. In addition, the segmented (joinpoint) trends introduced in this revision were fitted to central estimates, so their confidence intervals capture regression uncertainty around the fitted segments only, not GBD estimation uncertainty.

## 5. Conclusions

Integrating four GBD 2023 cause modules through three estimation interfaces into a single 29-node spectrum^1 2^ yields four conclusions. First, childhood respiratory infectious mortality totalled 965,330 deaths (95% uncertainty interval 680,096–1,342,437) at ages 0–19 years in 2023, with whooping cough (11.7%) and tuberculosis (9.1%) ranking second and third and COVID-19 fifth (5.5%). Second, the spectrum diversified as it shrank: Shannon H rose 16.1% and the effective number of causes nearly doubled (5.57 to 9.94). Third, pandemic perturbation separated causes into distinct phenotypes— whooping cough alone rebounded above 100% (+111.0%) while tuberculosis was wholly inertial. Fourth, 19 of 29 causes are poverty-locked in sub-Saharan Africa and South Asia. These contributions converge on one takeaway: the era in which one or two pathogens dominate childhood respiratory mortality has ended, and prevention portfolios must shift from single-pathogen vaccines towards platform interventions specified on an age-by-geography matrix. Future work should extend national resolution to the full annual series, validate share responses with interrupted time series around vaccine-introduction cohorts such as India’s 2017 PCV rollout, and extend the framework to incidence.

## Declarations

### Ethics approval and consent to participate

Not applicable. This study uses publicly available, de-identified, aggregate modelled estimates and was exempt from institutional ethics review.

### Consent for publication

Not applicable.

### Availability of data and materials

The datasets analyzed are publicly available from the Global Burden of Disease Results Tool (GBD 2023, release v8352; Institute for Health Metrics and Evaluation), the WHO/UNICEF Estimates of National Immunization Coverage (WUENIC), and the WHO Global Tuberculosis Report 2024. The analysis tables and scripts generated during this study are available from the corresponding author on reasonable request. The country-level dataset of pathogen-attributable deaths (204 countries, the full 29-cause spectrum (26 LRI aetiologies plus tuberculosis, pertussis, and COVID-19), five time points 1990–2023) is provided as Additional file 1 (03_Dataset.xlsx).

### Presentation

This work has not been presented at any scientific meeting.

### Authors’ contributions

SC conceived and designed the study; SC, HC and DL curated and analyzed the data; SC drafted the manuscript; HC, DL, YM, YZ and CS interpreted the results and critically revised the manuscript. All authors read and approved the final manuscript.

## Acknowledgements

The authors thank the Institute for Health Metrics and Evaluation and the Global Burden of Disease collaborator network for making the underlying estimates publicly available.

## Additional files

**Additional file 1: Country-level dataset (03_Dataset.xlsx).** Pathogen-attributable lower respiratory infection deaths at ages 0–19 years for 204 countries and 26 aetiologies at five time points (1990, 2010, 2019, 2021, 2023), with 95% uncertainty intervals, GLOBOCAN 2022 population denominators and crude death rates per 100,000 (26,520 country × aetiology × year rows, plus top-country rankings, largest 1990–2023 trend countries and pandemic-window country changes), extended to the full 29-cause spectrum by an age-band long-format sheet covering tuberculosis (5 time points), pertussis (5 time points) and COVID-19 (2021 and 2023 only; earlier time points not applicable), 9,792 country × cause × age × year rows.

**Additional file 2: Supplementary Tables S1–S6, S7–S10 and S11–S13, and Supplementary Figures S1–S2 (Supplementary_Materials.docx).** Five-anchor-year deaths for all 29 causes (S1); full perturbation–recovery phenotype matrix (S2); diversity time series with exclusion sensitivities (S3); composition at anchor years (S4); poverty-lock shares (S5); WUENIC dose–response tables (S6); country-level distribution of the 26 LRI aetiologies in 2023 (S7); country-level trends across five time points (S8); national-level Shannon diversity summary for 204 countries at five time points, globally and by super-region (S9); level versus first-difference correlations between diversity and total deaths (S10). Pandemic-window analyses against 2017–2019 natural-variability bands: depth gradient (S11), V-shape ranking of the 26 LRI aetiologies (S12), COVID-19 synchronicity (S13) and Figures S1–S2.

Additional file 3: Joinpoint segmented-trend results. Supplementary_Joinpoint_Spectrum29.csv — segment-specific annual percent changes with 95% confidence intervals, joinpoint locations and average annual percent changes for total 29-cause deaths, Shannon diversity and the six leading pathogen shares, global, ages 0–19 years, 1990–2023 — together with the fully reproducible analysis script (code/joinpoint_spectrum29.py).

