## Supplementary for "Diversifying deaths: the shifting spectrum of childhood respiratory infectious mortality, 1990–2023: a systematic analysis of the Global Burden of Disease Study 2023"

All estimates are drawn from the Global Burden of Disease Study 2023 (GBD 2023, release v8352), ages 0–19 years, metric Deaths (number), for a 29-node spectrum (26 lower respiratory infection etiologies plus tuberculosis, COVID-19, and whooping cough). Uncertainty intervals (UIs) are GBD-reported 95% UIs propagated by summing lower and upper bounds across strata; pooled UIs are therefore conservative. Values are shown with thousands separators; UIs are shown as (lower–upper).

Contents: Supplementary Tables S1–S6, S7–S10 and S11–S13, and Supplementary Figures S1–S2 (this file). Additional file 1: 03_Dataset.xlsx — country-level dataset of pathogen-attributable LRI deaths at five time points (1990, 2010, 2019, 2021, 2023) (204 countries × 26 aetiologies × 5 time points), with 95% uncertainty intervals, population denominators and crude death rates per 100,000 population aged 0–19 years (separate file).

**Supplementary Table S1. Deaths (95% UI) at five anchor years (1990, 2010, 2019, 2021, 2023) for all 29 causes, global, ages 0–19 years**

| Cause | 1990, deaths (95% UI) | 2010, deaths (95% UI) | 2019, deaths (95% UI) | 2021, deaths (95% UI) | 2023, deaths (95% UI) |
|---|---|---|---|---|---|
| Streptococcus pneumoniae | 974,495 (741,296–1,247,461) | 467,895 (354,039–609,455) | 267,916 (202,417–344,627) | 195,298 (140,346–265,897) | 227,976 (164,927–306,384) |
| Whooping cough | 277,598 (142,482–485,679) | 147,943 (85,356–244,066) | 125,586 (72,772–204,658) | 53,538 (30,037–85,953) | 112,954 (64,576–185,708) |
| Tuberculosis | 259,063 (174,175–372,311) | 138,600 (93,076–195,556) | 99,704 (68,956–143,734) | 95,299 (65,704–135,594) | 87,764 (57,779–124,912) |
| Klebsiella pneumoniae | 204,296 (155,649–264,583) | 102,963 (78,044–134,827) | 89,608 (66,958–116,693) | 71,866 (52,014–97,832) | 85,107 (61,665–115,061) |
| COVID-19 | 0 (0–0) | 0 (0–0) | 0 (0–0) | 147,942 (133,667–165,001) | 52,899 (47,275–59,183) |
| Pseudomonas aeruginosa | 102,142 (77,712–132,289) | 55,311 (41,748–71,684) | 53,800 (40,480–70,008) | 44,385 (32,464–61,274) | 52,329 (38,054–71,182) |

| | | | | | |
|---|---|---|---|---|---|
| Staphylococcus aureus | 56,614 (44,267–72,256) | 36,298 (27,984–46,932) | 37,990 (29,186–48,407) | 32,462 (23,994–43,449) | 38,016 (28,306–50,092) |
| Escherichia coli | 66,264 (50,420–85,185) | 37,888 (28,440–49,250) | 38,020 (28,905–48,772) | 31,436 (22,980–41,889) | 37,286 (27,198–49,596) |
| Other Mycobacterium species (non-TB, non-Leprosy) | 61,336 (41,283–90,935) | 34,513 (22,620–51,500) | 35,186 (23,808–50,407) | 29,937 (19,875–43,794) | 35,259 (23,236–52,283) |
| Influenza | 93,763 (71,299–120,680) | 48,883 (36,852–63,985) | 55,265 (40,691–71,959) | 20,646 (13,324–32,001) | 29,806 (20,398–43,781) |
| Respiratory syncytial virus | 93,448 (72,797–119,278) | 50,369 (38,283–64,123) | 54,480 (41,014–70,470) | 18,960 (12,323–29,125) | 28,052 (19,384–40,670) |
| Mycoplasma | 53,547 (40,085–69,818) | 28,359 (21,145–37,203) | 27,405 (20,451–36,027) | 22,725 (16,373–31,198) | 26,382 (18,984–36,012) |
| Group A Streptococcus | 44,520 (34,257–57,206) | 24,231 (18,673–31,088) | 20,131 (15,292–25,731) | 16,467 (12,108–22,360) | 19,333 (14,206–25,828) |
| Other gram-negative bacteria | 37,711 (28,680–48,712) | 20,206 (15,186–26,390) | 20,152 (14,914–26,396) | 16,391 (11,539–22,578) | 19,079 (13,870–25,918) |
| Haemophilus influenzae | 43,840 (33,982–56,166) | 22,632 (17,319–29,172) | 18,756 (14,156–24,295) | 15,025 (10,926–20,334) | 17,775 (12,876–23,974) |
| Other bacterial and viral pathogens | 33,121 (23,529–45,138) | 17,781 (12,429–24,311) | 16,418 (11,589–22,138) | 13,590 (9,604–18,859) | 16,173 (11,417–22,355) |
| Chlamydia spp | 37,233 (26,803–50,510) | 17,213 (12,061–23,589) | 15,308 (10,993–20,628) | 12,384 (8,556–17,316) | 14,663 (10,406–20,192) |
| Acinetobacter baumannii | 40,951 (30,724–54,059) | 19,738 (14,373–26,210) | 13,508 (9,988–17,911) | 10,294 (7,275–14,133) | 12,098 (8,530–16,501) |
| Aspergillus spp. | 22,021 (15,370–31,383) | 13,553 (9,716–18,716) | 12,196 (8,999–16,494) | 10,189 (7,198–14,141) | 11,781 (8,341–16,181) |
| Group B streptococcus | 20,640 (14,961–27,905) | 10,848 (7,736–15,070) | 11,182 (8,155–15,176) | 9,338 (6,615–13,140) | 11,110 (7,867–15,674) |
| Other fungi | 14,737 (10,515–20,222) | 8,227 (5,755–11,550) | 7,517 (5,238–10,279) | 6,516 (4,453–9,234) | 7,496 (5,133–10,584) |
| Other Klebsiella species | 8,612 (6,620–10,965) | 5,245 (3,891–6,802) | 5,765 (4,339–7,530) | 4,844 (3,581–6,572) | 5,690 (4,134–7,571) |

| | | | | | |
|---|---|---|---|---|---|
| Enterobacter spp | 8,630 (6,485–11,280) | 4,946 (3,735–6,436) | 4,958 (3,723–6,389) | 4,086 (3,020–5,562) | 4,870 (3,554–6,594) |
| Other Acinetobacter species | 8,358 (6,125–11,457) | 4,339 (3,103–5,952) | 4,287 (3,078–5,836) | 3,562 (2,467–5,070) | 4,143 (2,882–5,847) |
| Serratia spp. | 4,197 (3,173–5,484) | 2,492 (1,869–3,239) | 2,773 (2,088–3,656) | 2,348 (1,695–3,180) | 2,782 (2,028–3,793) |
| Citrobacter spp. | 2,403 (1,614–3,451) | 1,590 (1,077–2,269) | 1,630 (1,112–2,365) | 1,405 (941–2,049) | 1,666 (1,115–2,416) |
| Proteus spp. | 1,744 (1,255–2,359) | 1,118 (803–1,518) | 1,259 (936–1,716) | 1,126 (802–1,576) | 1,304 (943–1,813) |
| Morganella spp. | 1,929 (1,222–2,994) | 1,046 (666–1,600) | 972 (637–1,465) | 806 (515–1,269) | 942 (605–1,487) |
| Legionella spp | 276 (176–405) | 455 (314–625) | 562 (396–768) | 519 (358–726) | 594 (407–846) |
| **All 29 causes (total)** | **2,573,489 (1,856,957–3,500,170)** | **1,324,685 (956,292–1,803,118)** | **1,042,336 (751,273–1,414,534)** | **893,387 (654,755–1,211,106)** | **965,330 (680,096–1,342,437)** |

*Note: Each cell shows the point estimate followed by the 95% uncertainty interval (lower–upper). The 26 LRI etiologies are rei-level PAF-attributed deaths; tuberculosis, COVID-19, and whooping cough are independently modelled GBD causes. COVID-19 estimates are zero before 2020. The total row sums point estimates and UI bounds across all 29 nodes (conservative bound-summation propagation).*

### Supplementary Table S2. Full perturbation–recovery phenotype matrix for the 29-node spectrum, global, ages 0–19 years

| Cause | Type | Deaths 2019 | Deaths 2020 | Deaths 2021 | Deaths 2023 | Collapse 2019 to 2021, % | Rebound 2021 to 2023, % | Net change 2019 to 2023, % | Absolute rebound, deaths | Note |
|---|---|---|---|---|---|---|---|---|---|---|
| Acinetobacter baumannii | Bacterium | 13,508 | 11,481 | 10,294 | 12,098 | -23.8 | 17.5 | -10.4 | 1,804 | – |
| Aspergillus spp. | Fungus | 12,196 | 10,833 | 10,189 | 11,781 | -16.5 | 15.6 | -3.4 | 1,592 | – |
| COVID-19 | Pandemic-driven | 0 | 99,113 | 147,942 | 52,899 | – | – | – | – | 2019 baseline = 0 |

| | | | | | | | | | | |
|---|---|---|---|---|---|---|---|---|---|---|
| | (new virus) | | | | | | | | | (pathogen absent); peak 2021, decline through 2023 |
| Chlamydia spp | Bacterium | 15,308 | 13,434 | 12,384 | 14,663 | -19.1 | 18.4 | -4.2 | 2,279 | – |
| Citrobacter spp. | Bacterium | 1,630 | 1,466 | 1,405 | 1,666 | -13.8 | 18.5 | 2.2 | 261 | – |
| Enterobacter spp | Bacterium | 4,958 | 4,371 | 4,086 | 4,870 | -17.6 | 19.2 | -1.8 | 785 | – |
| Escherichia coli | Bacterium | 38,020 | 33,587 | 31,436 | 37,286 | -17.3 | 18.6 | -1.9 | 5,850 | – |
| Group A Streptococcus | Bacterium | 20,131 | 17,561 | 16,467 | 19,333 | -18.2 | 17.4 | -4.0 | 2,866 | – |
| Group B streptococcus | Bacterium | 11,182 | 9,958 | 9,338 | 11,110 | -16.5 | 19.0 | -0.6 | 1,771 | – |
| Haemophilus influenzae | Bacterium | 18,756 | 16,273 | 15,025 | 17,775 | -19.9 | 18.3 | -5.2 | 2,750 | – |
| Influenza | Virus | 55,265 | 26,388 | 20,646 | 29,806 | -62.6 | 44.4 | -46.1 | 9,160 | – |
| Klebsiella pneumoniae | Bacterium | 89,608 | 78,065 | 71,866 | 85,107 | -19.8 | 18.4 | -5.0 | 13,241 | – |
| Legionella spp | Bacterium | 562 | 507 | 519 | 594 | -7.7 | 14.4 | 5.6 | 75 | – |

| | | | | | | | | | | |
|---|---|---|---|---|---|---|---|---|---|---|
| Morganella spp. | Bacterium | 972 | 864 | 806 | 942 | -17.1 | 16.9 | -3.1 | 136 | – |
| Mycoplasma | Bacterium | 27,405 | 24,338 | 22,725 | 26,382 | -17.1 | 16.1 | -3.7 | 3,657 | – |
| Other Acinetobacter species | Bacterium | 4,287 | 3,813 | 3,562 | 4,143 | -16.9 | 16.3 | -3.4 | 581 | – |
| Other Klebsiella species | Bacterium | 5,765 | 5,139 | 4,844 | 5,690 | -16.0 | 17.5 | -1.3 | 846 | – |
| Other Mycobacterium species (non-TB, non-Leprosy) | Bacterium | 35,186 | 31,635 | 29,937 | 35,259 | -14.9 | 17.8 | 0.2 | 5,321 | – |
| Other bacterial and viral pathogens | Other (mixed bacterial/viral) | 16,418 | 14,533 | 13,590 | 16,173 | -17.2 | 19.0 | -1.5 | 2,583 | – |
| Other fungi | Fungus | 7,517 | 6,821 | 6,516 | 7,496 | -13.3 | 15.0 | -0.3 | 979 | – |
| Other gram-negative bacteria | Bacterium | 20,152 | 17,750 | 16,391 | 19,079 | -18.7 | 16.4 | -5.3 | 2,688 | – |
| Proteus spp. | Bacterium | 1,259 | 1,161 | 1,126 | 1,304 | -10.6 | 15.8 | 3.6 | 178 | – |
| Pseudomonas aeruginosa | Bacterium | 53,800 | 47,550 | 44,385 | 52,329 | -17.5 | 17.9 | -2.7 | 7,944 | – |

| | | | | | | | | | | |
|---|---|---|---|---|---|---|---|---|---|---|
| Respiratory syncytial virus | Virus | 54,480 | 25,079 | 18,960 | 28,052 | -65.2 | 48.0 | -48.5 | 9,092 | – |
| Serratia spp. | Bacterium | 2,773 | 2,482 | 2,348 | 2,782 | -15.3 | 18.5 | 0.3 | 434 | – |
| Staphylococcus aureus | Bacterium | 37,990 | 34,047 | 32,462 | 38,016 | -14.6 | 17.1 | 0.1 | 5,555 | – |
| Streptococcus pneumoniae | Bacterium | 267,916 | 221,719 | 195,298 | 227,976 | -27.1 | 16.7 | -14.9 | 32,678 | – |
| Tuberculosis | Bacterium | 99,704 | 98,077 | 95,299 | 87,764 | -4.4 | -7.9 | -12.0 | -7,535 | – |
| Whooping cough | Bacterium | 125,586 | 71,253 | 53,538 | 112,954 | -57.4 | 111.0 | -10.1 | 59,416 | – |

*Note: Collapse is the percentage change from 2019 to 2021; rebound is the percentage recovery from 2021 to 2023; net change is 2023 versus 2019; absolute rebound is the 2023-minus-trough difference in deaths. Metrics are undefined (shown as –) for COVID-19 because the 2019 baseline is zero. Relative and absolute rebound ranks and collapse-depth ranks are available in the underlying CSV (x2_pandemic_phenotype_matrix_29.csv). Legionella is the single exception to the 2021 trough convention (its true trough was 2020: collapse −9.8%, absolute rebound 87 deaths; the table reports the 2021-window values −7.7% and 75).*

**Supplementary Table S3. Diversity time series 1990–2023 for the global 0–19 death spectrum: Shannon diversity (H), Herfindahl–Hirschman index (HHI), and effective number of causes (effN), for the full 29-node spectrum and two 28-node sensitivity series**

| Year | All 29 causes | | | Excluding COVID-19 (28) | | | Excluding tuberculosis (28) | | |
|---|---|---|---|---|---|---|---|---|---|
| | H | HHI | effN | H | HHI | effN | H | HHI | effN |
| 1990 | 2.336 | 0.179 | 5.57 | 2.336 | 0.179 | 5.57 | 2.234 | 0.209 | 4.78 |
| 1991 | 2.338 | 0.179 | 5.58 | 2.338 | 0.179 | 5.58 | 2.237 | 0.209 | 4.78 |
| 1992 | 2.340 | 0.179 | 5.59 | 2.340 | 0.179 | 5.59 | 2.239 | 0.209 | 4.78 |
| 1993 | 2.342 | 0.179 | 5.60 | 2.342 | 0.179 | 5.60 | 2.242 | 0.209 | 4.78 |
| 1994 | 2.343 | 0.179 | 5.60 | 2.343 | 0.179 | 5.60 | 2.243 | 0.209 | 4.78 |
| 1995 | 2.344 | 0.178 | 5.62 | 2.344 | 0.178 | 5.62 | 2.243 | 0.209 | 4.79 |

| | | | | | | | | | |
|---|---|---|---|---|---|---|---|---|---|
| 1996 | 2.344 | 0.178 | 5.63 | 2.344 | 0.178 | 5.63 | 2.245 | 0.209 | 4.79 |
| 1997 | 2.346 | 0.177 | 5.66 | 2.346 | 0.177 | 5.66 | 2.246 | 0.208 | 4.81 |
| 1998 | 2.347 | 0.176 | 5.68 | 2.347 | 0.176 | 5.68 | 2.248 | 0.207 | 4.83 |
| 1999 | 2.349 | 0.175 | 5.70 | 2.349 | 0.175 | 5.70 | 2.250 | 0.206 | 4.85 |
| 2000 | 2.351 | 0.175 | 5.73 | 2.351 | 0.175 | 5.73 | 2.252 | 0.205 | 4.87 |
| 2001 | 2.354 | 0.174 | 5.76 | 2.354 | 0.174 | 5.76 | 2.256 | 0.204 | 4.90 |
| 2002 | 2.357 | 0.173 | 5.77 | 2.357 | 0.173 | 5.77 | 2.259 | 0.203 | 4.92 |
| 2003 | 2.361 | 0.172 | 5.80 | 2.361 | 0.172 | 5.80 | 2.262 | 0.202 | 4.96 |
| 2004 | 2.364 | 0.172 | 5.82 | 2.364 | 0.172 | 5.82 | 2.266 | 0.201 | 4.97 |
| 2005 | 2.367 | 0.171 | 5.84 | 2.367 | 0.171 | 5.84 | 2.269 | 0.200 | 4.99 |
| 2006 | 2.369 | 0.171 | 5.86 | 2.369 | 0.171 | 5.86 | 2.272 | 0.199 | 5.01 |
| 2007 | 2.373 | 0.170 | 5.90 | 2.373 | 0.170 | 5.90 | 2.276 | 0.198 | 5.05 |
| 2008 | 2.380 | 0.168 | 5.94 | 2.380 | 0.168 | 5.94 | 2.284 | 0.197 | 5.09 |
| 2009 | 2.390 | 0.166 | 6.04 | 2.390 | 0.166 | 6.04 | 2.295 | 0.193 | 5.18 |
| 2010 | 2.400 | 0.163 | 6.13 | 2.400 | 0.163 | 6.13 | 2.307 | 0.190 | 5.27 |
| 2011 | 2.415 | 0.160 | 6.26 | 2.415 | 0.160 | 6.26 | 2.323 | 0.185 | 5.39 |
| 2012 | 2.435 | 0.155 | 6.46 | 2.435 | 0.155 | 6.46 | 2.345 | 0.179 | 5.58 |
| 2013 | 2.455 | 0.150 | 6.67 | 2.455 | 0.150 | 6.67 | 2.367 | 0.173 | 5.78 |
| 2014 | 2.478 | 0.144 | 6.93 | 2.478 | 0.144 | 6.93 | 2.393 | 0.166 | 6.03 |
| 2015 | 2.508 | 0.137 | 7.28 | 2.508 | 0.137 | 7.28 | 2.425 | 0.157 | 6.37 |
| 2016 | 2.539 | 0.130 | 7.68 | 2.539 | 0.130 | 7.68 | 2.459 | 0.148 | 6.75 |
| 2017 | 2.567 | 0.124 | 8.09 | 2.567 | 0.124 | 8.09 | 2.490 | 0.140 | 7.14 |
| 2018 | 2.595 | 0.118 | 8.50 | 2.595 | 0.118 | 8.50 | 2.520 | 0.133 | 7.54 |
| 2019 | 2.620 | 0.112 | 8.94 | 2.620 | 0.112 | 8.94 | 2.549 | 0.126 | 7.96 |
| 2020 | 2.685 | 0.103 | 9.70 | 2.626 | 0.115 | 8.71 | 2.625 | 0.115 | 8.71 |
| 2021 | 2.648 | 0.106 | 9.41 | 2.636 | 0.113 | 8.82 | 2.584 | 0.119 | 8.41 |
| 2022 | 2.702 | 0.100 | 9.96 | 2.633 | 0.110 | 9.11 | 2.640 | 0.112 | 8.96 |

| | | | | | | | | | |
|---|---|---|---|---|---|---|---|---|---|
| 2023 | 2.711 | 0.101 | 9.94 | 2.644 | 0.109 | 9.15 | 2.647 | 0.112 | 8.95 |

*Note: H = –Σ p ln p over cause shares; HHI = Σ p²; effN = 1/HHI. Sensitivity series exclude COVID-19 (28 nodes) or tuberculosis (28 nodes) and are renormalized to their respective yearly totals. The entry of COVID-19 in 2020 is a structural break in the 29-node series.*

**Supplementary Table S4. Composition of the 29-node spectrum at anchor years (1990, 2010, 2019, 2023): rank and share of the yearly 29-node total, global, ages 0–19 years**

| Cause | Rank | | | | Share of 29-node total, % | | | |
|---|---|---|---|---|---|---|---|---|
| | 1990 | 2010 | 2019 | 2023 | 1990 | 2010 | 2019 | 2023 |
| Streptococcus pneumoniae | 1 | 1 | 1 | 1 | 37.9 | 35.3 | 25.7 | 23.6 |
| Whooping cough | 2 | 2 | 2 | 2 | 10.8 | 11.2 | 12.0 | 11.7 |
| Tuberculosis | 3 | 3 | 3 | 3 | 10.1 | 10.5 | 9.6 | 9.1 |
| Klebsiella pneumoniae | 4 | 4 | 4 | 4 | 7.9 | 7.8 | 8.6 | 8.8 |
| COVID-19 | 29 | 29 | 29 | 5 | 0.0 | 0.0 | 0.0 | 5.5 |
| Pseudomonas aeruginosa | 5 | 5 | 7 | 6 | 4.0 | 4.2 | 5.2 | 5.4 |
| Staphylococcus aureus | 10 | 9 | 9 | 7 | 2.2 | 2.7 | 3.6 | 3.9 |
| Escherichia coli | 8 | 8 | 8 | 8 | 2.6 | 2.9 | 3.6 | 3.9 |
| Other Mycobacterium species (non-TB, non-Leprosy) | 9 | 10 | 10 | 9 | 2.4 | 2.6 | 3.4 | 3.7 |
| Influenza | 6 | 7 | 5 | 10 | 3.6 | 3.7 | 5.3 | 3.1 |

| | | | | | | | | |
|---|---|---|---|---|---|---|---|---|
| Respiratory syncytial virus | 7 | 6 | 6 | 11 | 3.6 | 3.8 | 5.2 | 2.9 |
| Mycoplasma | 11 | 11 | 11 | 12 | 2.1 | 2.1 | 2.6 | 2.7 |
| Group A Streptococcus | 12 | 12 | 13 | 13 | 1.7 | 1.8 | 1.9 | 2.0 |
| Other gram-negative bacteria | 15 | 14 | 12 | 14 | 1.5 | 1.5 | 1.9 | 2.0 |
| Haemophilus influenzae | 13 | 13 | 14 | 15 | 1.7 | 1.7 | 1.8 | 1.8 |
| Other bacterial and viral pathogens | 17 | 16 | 15 | 16 | 1.3 | 1.3 | 1.6 | 1.7 |
| Chlamydia spp | 16 | 17 | 16 | 17 | 1.4 | 1.3 | 1.5 | 1.5 |
| Acinetobacter baumannii | 14 | 15 | 17 | 18 | 1.6 | 1.5 | 1.3 | 1.3 |
| Aspergillus spp. | 18 | 18 | 18 | 19 | 0.9 | 1.0 | 1.2 | 1.2 |
| Group B streptococcus | 19 | 19 | 19 | 20 | 0.8 | 0.8 | 1.1 | 1.2 |
| Other fungi | 20 | 20 | 20 | 21 | 0.6 | 0.6 | 0.7 | 0.8 |
| Other Klebsiella species | 22 | 21 | 21 | 22 | 0.3 | 0.4 | 0.6 | 0.6 |
| Enteroba | 21 | 22 | 22 | 23 | 0.3 | 0.4 | 0.5 | 0.5 |

| | | | | | | | | |
|---|---|---|---|---|---|---|---|---|
| cter spp | | | | | | | | |
| Other Acinetobacter species | 23 | 23 | 23 | 24 | 0.3 | 0.3 | 0.4 | 0.4 |
| Serratia spp. | 24 | 24 | 24 | 25 | 0.2 | 0.2 | 0.3 | 0.3 |
| Citrobacter spp. | 25 | 25 | 25 | 26 | 0.1 | 0.1 | 0.2 | 0.2 |
| Proteus spp. | 27 | 26 | 26 | 27 | 0.1 | 0.1 | 0.1 | 0.1 |
| Morganella spp. | 26 | 27 | 27 | 28 | 0.1 | 0.1 | 0.1 | 0.1 |
| Legionella spp | 28 | 28 | 28 | 29 | 0.0 | 0.0 | 0.1 | 0.1 |

*Note: Shares are percentages of the yearly all-cause 29-node total; ranks are ordered by deaths within each year. COVID-19 contributed zero deaths before 2020 and is unranked in 1990–2019 in the underlying CSV; its rank cells for those years reflect the zero-death ordering.*

### Supplementary Table S5. Poverty-lock shares in 2023: combined sub-Saharan Africa and South Asia share of each node's deaths, 29 nodes, ages 0–19 years

| Cause | Type | SSA + South Asia share 2023, % | Poverty-locked |
|---|---|---|---|
| Tuberculosis | Bacterium | 87.1 | Yes |
| Chlamydia spp | Bacterium | 86.0 | Yes |
| Acinetobacter baumannii | Bacterium | 84.3 | Yes |
| Group B streptococcus | Bacterium | 83.8 | Yes |
| Klebsiella pneumoniae | Bacterium | 83.6 | Yes |
| Morganella spp. | Bacterium | 83.0 | Yes |
| Other bacterial and viral pathogens | Other (mixed bacterial/viral) | 83.0 | Yes |
| Other gram-negative bacteria | Bacterium | 82.9 | Yes |
| Whooping cough | Bacterium | 82.7 | Yes |

| | | | |
|---|---|---|---|
| Other Acinetobacter species | Bacterium | 82.6 | Yes |
| Other Mycobacterium species (non-TB, non-Leprosy) | Bacterium | 82.3 | Yes |
| Pseudomonas aeruginosa | Bacterium | 82.2 | Yes |
| Streptococcus pneumoniae | Bacterium | 81.7 | Yes |
| Mycoplasma | Bacterium | 81.6 | Yes |
| Influenza | Virus | 81.1 | Yes |
| Escherichia coli | Bacterium | 80.9 | Yes |
| Enterobacter spp | Bacterium | 80.9 | Yes |
| Serratia spp. | Bacterium | 80.7 | Yes |
| Haemophilus influenzae | Bacterium | 80.6 | Yes |
| Other fungi | Fungus | 79.3 | No |
| Respiratory syncytial virus | Virus | 79.3 | No |
| Other Klebsiella species | Bacterium | 79.3 | No |
| Proteus spp. | Bacterium | 78.4 | No |
| Group A Streptococcus | Bacterium | 77.0 | No |
| Aspergillus spp. | Fungus | 76.8 | No |
| Staphylococcus aureus | Bacterium | 76.6 | No |
| Citrobacter spp. | Bacterium | 76.1 | No |
| COVID-19 | Virus | 54.0 | No |
| Legionella spp | Bacterium | 29.8 | No |

*Note: Nodes whose combined sub-Saharan Africa (SSA) and South Asia share exceeds the all-cause reference line of 80.59% (the two regions' joint share of all 29-node deaths in 2023) are classified as poverty-locked (19 of 29 nodes). The classification is denominator-relative: the corresponding 26-pathogen reference was 81.44%.*

**Supplementary Table S6. WUENIC dose–response: super-region PCV3 and Hib3 coverage (2015, three calibers) versus 2010 to 2023 change in the target cause's share of the 29-node super-region total, ages 0–19 years**

| Vaccine | GBD | Coverag | Coverag | Coverag | Countrie | Introduc | Share | Share | Share |
|---|---|---|---|---|---|---|---|---|---|

| | super-region | e 2015, median, all countries, % | e 2015, mean, introduced countries, % | e 2015, population-weighted mean, % | s, n | ed by 2015, n | 2010, % | 2023, % | change 2010 to 2023, pp |
|---|---|---|---|---|---|---|---|---|---|
| PCV3 | Central Europe, Eastern Europe, and Central Asia | 0.0 | 72.1 | 12.2 | 29 | 11 | 43.1 | 22.6 | -20.6 |
| PCV3 | High-income | 91.0 | 90.4 | 85.6 | 35 | 28 | 29.9 | 13.7 | -16.2 |
| PCV3 | Latin America and Caribbean | 83.5 | 84.8 | 82.8 | 30 | 19 | 36.1 | 22.6 | -13.4 |
| PCV3 | North Africa and Middle East | 71.0 | 89.9 | 41.9 | 21 | 13 | 37.5 | 24.6 | -12.9 |
| PCV3 | South Asia | 5.0 | 44.3 | 14.5 | 5 | 3 | 35.9 | 27.0 | -8.8 |
| PCV3 | Southeast Asia, East Asia, and Oceania | 0.0 | 66.7 | 1.9 | 29 | 11 | 35.7 | 21.1 | -14.5 |
| PCV3 | Sub-Saharan Africa | 71.5 | 72.8 | 61.6 | 46 | 37 | 34.3 | 22.6 | -11.7 |
| Hib3 | Central Europe, Eastern Europe, and Central Asia | 93.0 | 87.7 | 52.7 | 29 | 28 | 2.3 | 2.7 | 0.4 |
| Hib3 | High- | 96.0 | 95.3 | 94.7 | 35 | 35 | 2.2 | 2.4 | 0.1 |

| | | | | | | | | | |
|---|---|---|---|---|---|---|---|---|---|
| | income | | | | | | | | |
| Hib3 | Latin America and Caribbean | 92.0 | 89.7 | 90.1 | 30 | 30 | 1.7 | 2.2 | 0.5 |
| Hib3 | North Africa and Middle East | 98.0 | 90.1 | 89.2 | 21 | 21 | 2.0 | 2.0 | -0.0 |
| Hib3 | South Asia | 91.0 | 81.0 | 54.0 | 5 | 5 | 2.0 | 2.2 | 0.3 |
| Hib3 | Southeast Asia, East Asia, and Oceania | 89.0 | 87.4 | 26.0 | 29 | 27 | 1.9 | 1.4 | -0.5 |
| Hib3 | Sub-Saharan Africa | 84.0 | 77.6 | 70.0 | 46 | 46 | 1.4 | 1.7 | 0.2 |

*Note: Coverage is the 2015 WUENIC estimate (WHO/UNICEF Estimates of National Immunization Coverage) aggregated to GBD super-regions. A WUENIC value of 0 denotes non-introduction of the vaccine into the national programme, not true zero coverage; the "introduced countries" caliber excludes such zeros. Shares refer to the vaccine-target cause (Streptococcus pneumoniae for PCV3; Haemophilus influenzae for Hib3) as a percentage of the 29-node super-region total. Spearman rank correlation of the median-all caliber versus share change: PCV3 ρ = +0.108, p = 0.818; Hib3 ρ = −0.036, p = 0.939 (n = 7 super-regions each; hypothesis-generating).*

**Supplementary Table S7. Country-level distribution of the 26 LRI aetiologies, 2023: global deaths and the top three countries by deaths with their shares of each aetiology's global total, ages 0–19 years**

| Aetiology | Global deaths 2023 (95% UI) | Top three countries by deaths: deaths (share of aetiology global total, %) |
|---|---|---|
| Acinetobacter baumannii | 12,098 (8,530–16,501) | India 2,781 (23.0%); Nigeria 2,030 (16.8%); Democratic Republic of the Congo 520 (4.3%) |
| Aspergillus spp. | 11,781 (8,341–16,181) | India 2,282 (19.4%); Nigeria 1,798 (15.3%); Pakistan 517 (4.4%) |

| | | |
|---|---|---|
| Chlamydia spp | 14,663 (10,406–20,192) | India 2,651 (18.1%); Nigeria 2,309 (15.8%); Pakistan 904 (6.2%) |
| Citrobacter spp. | 1,666 (1,115–2,416) | India 422 (25.3%); Nigeria 204 (12.3%); Pakistan 70 (4.2%) |
| Enterobacter spp | 4,870 (3,554–6,594) | India 1,145 (23.5%); Nigeria 670 (13.8%); Pakistan 263 (5.4%) |
| Escherichia coli | 37,286 (27,198–49,596) | India 8,143 (21.9%); Nigeria 5,347 (14.3%); Pakistan 2,131 (5.7%) |
| Group A Streptococcus | 19,333 (14,206–25,828) | India 3,631 (18.8%); Nigeria 3,044 (15.8%); Pakistan 1,129 (5.8%) |
| Group B streptococcus | 11,110 (7,867–15,674) | India 2,246 (20.2%); Nigeria 1,546 (13.9%); Pakistan 676 (6.1%) |
| Haemophilus influenzae | 17,775 (12,876–23,974) | India 3,836 (21.6%); Nigeria 2,967 (16.7%); Pakistan 977 (5.5%) |
| Influenza | 29,806 (20,398–43,781) | Nigeria 5,465 (18.3%); India 3,185 (10.7%); Democratic Republic of the Congo 2,021 (6.8%) |
| Klebsiella pneumoniae | 85,107 (61,665–115,061) | India 17,558 (20.6%); Nigeria 13,187 (15.5%); Pakistan 4,782 (5.6%) |
| Legionella spp | 594 (407–846) | China 118 (19.9%); India 76 (12.9%); United States of America 26 (4.3%) |
| Morganella spp. | 942 (605–1,487) | India 179 (19.0%); Nigeria 144 (15.3%); Pakistan 49 (5.3%) |
| Mycoplasma | 26,382 (18,984–36,012) | Nigeria 4,469 (16.9%); India 4,186 (15.9%); Pakistan 1,563 (5.9%) |
| Other Acinetobacter species | 4,143 (2,882–5,847) | Nigeria 695 (16.8%); India 633 (15.3%); Pakistan 261 (6.3%) |
| Other Klebsiella species | 5,690 (4,134–7,571) | India 1,112 (19.5%); Nigeria 839 (14.8%); Pakistan 341 (6.0%) |
| Other Mycobacterium species (non-TB, non-Leprosy) | 35,259 (23,236–52,283) | India 7,099 (20.1%); Nigeria 4,929 (14.0%); Pakistan 1,881 (5.3%) |
| Other bacterial and viral pathogens | 16,173 (11,417–22,355) | India 3,752 (23.2%); Nigeria 2,270 (14.0%); Pakistan 819 (5.1%) |
| Other fungi | 7,496 (5,133–10,584) | Nigeria 1,251 (16.7%); India 1,167 (15.6%); Pakistan 380 (5.1%) |
| Other gram-negative bacteria | 19,079 (13,870–25,918) | Nigeria 3,188 (16.7%); India 3,064 |

| | | |
|---|---|---|
| | | (16.1%); Pakistan 1,166 (6.1%) |
| Proteus spp. | 1,304 (943–1,813) | India 224 (17.2%); Nigeria 171 (13.2%); Pakistan 64 (4.9%) |
| Pseudomonas aeruginosa | 52,329 (38,054–71,182) | India 10,100 (19.3%); Nigeria 8,083 (15.5%); Pakistan 3,156 (6.0%) |
| Respiratory syncytial virus | 28,052 (19,384–40,670) | Nigeria 4,973 (17.7%); India 3,917 (14.0%); Democratic Republic of the Congo 1,680 (6.0%) |
| Serratia spp. | 2,782 (2,028–3,793) | India 580 (20.8%); Nigeria 372 (13.4%); Pakistan 161 (5.8%) |
| Staphylococcus aureus | 38,016 (28,306–50,092) | India 7,501 (19.7%); Nigeria 5,062 (13.3%); Pakistan 2,004 (5.3%) |
| Streptococcus pneumoniae | 227,976 (164,927–306,384) | India 51,724 (22.7%); Nigeria 41,447 (18.2%); Democratic Republic of the Congo 9,988 (4.4%) |

*Note: Deaths are GBD 2023 rei-level PAF-attributed lower respiratory infection (LRI) deaths at ages 0–19 years, summed across the four age bands (<5, 5–9, 10–14, 15–19); 95% uncertainty intervals (UIs) are propagated by summing bounds. Share is the country's percentage of the aetiology's 2023 global total (sum over 204 countries); country names are GBD location names. The full 204 × 26 matrix—including ISO3 codes, GBD super-regions, GLOBOCAN 2022 population denominators (ages 0–19) and crude death rates per 100,000—is provided in Additional file 1 (03_Dataset.xlsx).*

**Supplementary Table S8. Country-level trends across five time points: global deaths (ages 0–19 years) attributable to each of the 26 LRI aetiologies at 1990, 2010, 2019, 2021 and 2023, and percentage change from 1990 to 2023**

| Aetiology | 1990 | 2010 | 2019 | 2021 | 2023 | Change 1990–2023 (%) |
|---|---|---|---|---|---|---|
| Streptococcus pneumoniae | 974,495 | 467,895 | 267,916 | 195,298 | 227,976 | -76.6 |
| Klebsiella pneumoniae | 204,296 | 102,963 | 89,608 | 71,866 | 85,107 | -58.3 |
| Pseudomonas aeruginosa | 102,142 | 55,311 | 53,800 | 44,385 | 52,329 | -48.8 |
| Staphylococcus aureus | 56,614 | 36,298 | 37,990 | 32,462 | 38,016 | -32.8 |
| Escherichia | 66,264 | 37,888 | 38,020 | 31,436 | 37,286 | -43.7 |

| | | | | | | |
|---|---|---|---|---|---|---|
| coli | | | | | | |
| Other Mycobacterium species (non-TB, non-Leprosy) | 61,336 | 34,513 | 35,186 | 29,937 | 35,259 | -42.5 |
| Influenza | 93,763 | 48,883 | 55,265 | 20,646 | 29,806 | -68.2 |
| Respiratory syncytial virus | 93,448 | 50,369 | 54,480 | 18,960 | 28,052 | -70.0 |
| Mycoplasma | 53,547 | 28,359 | 27,405 | 22,725 | 26,382 | -50.7 |
| Group A Streptococcus | 44,520 | 24,231 | 20,131 | 16,467 | 19,333 | -56.6 |
| Other gram-negative bacteria | 37,711 | 20,206 | 20,152 | 16,391 | 19,079 | -49.4 |
| Haemophilus influenzae | 43,840 | 22,632 | 18,756 | 15,025 | 17,775 | -59.5 |
| Other bacterial and viral pathogens | 33,121 | 17,781 | 16,418 | 13,590 | 16,173 | -51.2 |
| Chlamydia spp | 37,233 | 17,213 | 15,308 | 12,384 | 14,663 | -60.6 |
| Acinetobacter baumannii | 40,951 | 19,738 | 13,508 | 10,294 | 12,098 | -70.5 |
| Aspergillus spp. | 22,021 | 13,553 | 12,196 | 10,189 | 11,781 | -46.5 |
| Group B streptococcus | 20,640 | 10,848 | 11,182 | 9,338 | 11,110 | -46.2 |
| Other fungi | 14,737 | 8,227 | 7,517 | 6,516 | 7,496 | -49.1 |
| Other Klebsiella species | 8,612 | 5,245 | 5,765 | 4,844 | 5,690 | -33.9 |
| Enterobacter spp | 8,630 | 4,946 | 4,958 | 4,086 | 4,870 | -43.6 |

| | | | | | | |
|---|---|---|---|---|---|---|
| Other Acinetobacter species | 8,358 | 4,339 | 4,287 | 3,562 | 4,143 | -50.4 |
| Serratia spp. | 4,197 | 2,492 | 2,773 | 2,348 | 2,782 | -33.7 |
| Citrobacter spp. | 2,403 | 1,590 | 1,630 | 1,405 | 1,666 | -30.7 |
| Proteus spp. | 1,744 | 1,118 | 1,259 | 1,126 | 1,304 | -25.2 |
| Morganella spp. | 1,929 | 1,046 | 972 | 806 | 942 | -51.2 |
| Legionella spp | 276 | 455 | 562 | 519 | 594 | +115.0 |
| All 26 LRI aetiologies | 2,036,827 | 1,038,142 | 817,047 | 596,607 | 711,713 | -65.1 |

Note: Deaths are GBD 2023 rei-level PAF-attributed lower respiratory infection (LRI) deaths at ages 0–19 years, summed over the 204-country panel for each aetiology and time point; country-level sums reproduce the GBD 2023 global modelled values used in the manuscript within a maximum absolute relative difference of 0.087%. Change is the percentage difference between the 2023 and 1990 levels. The full country × aetiology × year matrix (26,520 rows; 204 countries × 26 aetiologies × 5 time points), the countries with the largest 1990–2023 absolute increases and decreases per aetiology, and country-level pandemic-window (2019–2021–2023) percentage changes are provided in Additional file 1 (03_Dataset.xlsx).

**Supplementary Table S9. National-level Shannon diversity of the 26-aetiology LRI mortality spectrum across 204 countries and territories at five time points (1990, 2010, 2019, 2021, 2023), globally and by GBD super-region, ages 0–19 years**

| GBD super-region | Countries, n | Median H 1990 | Median H 2010 | Median H 2019 | Median H 2021 | Median H 2023 | H rose 1990 to 2023, % | H fell 2019 to 2021, % | H recovered 2021 to 2023, % |
|---|---|---|---|---|---|---|---|---|---|
| Central Europe, Eastern Europe, and Central Asia | 29 | 2.15 | 2.26 | 2.54 | 2.49 | 2.61 | 100.0 | 86.2 | 96.6 |
| High-income | 36 | 2.18 | 2.53 | 2.73 | 2.68 | 2.75 | 100.0 | 91.7 | 94.4 |
| Latin America and Caribbe | 33 | 2.09 | 2.35 | 2.66 | 2.58 | 2.63 | 100.0 | 90.9 | 90.9 |

| | | | | | | | | | |
|---|---|---|---|---|---|---|---|---|---|
| an | | | | | | | | | |
| North Africa and Middle East | 21 | 2.11 | 2.31 | 2.66 | 2.64 | 2.69 | 100.0 | 42.9 | 90.5 |
| South Asia | 5 | 2.20 | 2.22 | 2.58 | 2.60 | 2.64 | 100.0 | 0.0 | 60.0 |
| Southeast Asia, East Asia, and Oceania | 34 | 2.07 | 2.22 | 2.48 | 2.39 | 2.46 | 100.0 | 97.1 | 100.0 |
| Sub-Saharan Africa | 46 | 2.07 | 2.20 | 2.55 | 2.57 | 2.57 | 97.8 | 30.4 | 60.9 |
| **Global (all 204 countries)** | **204** | **2.11** | **2.30** | **2.59** | **2.57** | **2.62** | **99.5** | **70.6** | **86.3** |

*Note: H is the Shannon diversity index (−Σ p ln p) computed over the shares of the 26 rei-level PAF-attributed LRI aetiologies within each country-year (ages 0–19 years, point estimates; the four age bands are summed before shares are computed). Entries are medians across countries within each super-region; the Global row pools all 204 countries. "H rose 1990 to 2023" is the percentage of countries with higher H in 2023 than in 1990 (sole exception worldwide: Somalia, ΔH = −0.006); "H fell 2019 to 2021" and "H recovered 2021 to 2023" are the corresponding pandemic-window percentages. Source: Additional file 1 (03_Dataset.xlsx).*

#### Supplementary Table S10. Level versus first-difference correlations between annual diversity and annual total deaths, global ages 0–19 years, 1990–2023 (34 annual observations; 33 first differences).

| Diversity series (vs annual total deaths) | Levels: Spearman ρ (p) | Levels: Pearson r (p) | First differences: Spearman ρ (p) | First differences: Pearson r (p) |
|---|---|---|---|---|
| 29-node Shannon H | −0.997 ($9.6 \times 10^{-38}$) | −0.810 ($6.6 \times 10^{-9}$) | +0.487 (0.004) | +0.221 (0.216) |
| 26-node Shannon H (sensitivity) | −0.997 ($9.6 \times 10^{-38}$) | −0.840 ($5.4 \times 10^{-10}$) | +0.561 (<0.001) | +0.331 (0.060) |
| 29-node effN (1/HHI) | −0.998 ($1.7 \times 10^{-39}$) | −0.779 ($5.8 \times 10^{-8}$) | +0.319 (0.070) | +0.153 (0.395) |
| 26-node effN (1/HHI, sensitivity) | −0.996 ($9.6 \times 10^{-36}$) | −0.796 ($1.8 \times 10^{-8}$) | +0.641 (<0.001) | +0.457 (0.008) |

*Note: Level correlations relate annual diversity to annual total deaths; first-difference correlations relate year-to-year changes (Δdiversity versus Δdeaths, n = 33). effN = 1/HHI is the effective number of causes. The 26-node sensitivity series excludes tuberculosis, pertussis and COVID-19. The near-perfect inverse level correlations do not survive first-differencing: differenced correlations flip sign (positive) and, for the headline 29-node Shannon series, are non-significant by Pearson (p = 0.216), indicating that the level association is carried by the shared secular trend rather than by year-to-year inverse coupling. p values are two-sided.*

### Pandemic-window dynamics against natural variability (2017–2023)

Methods. For each of the 26 LRI aetiologies, global deaths at ages 0–19 years were assembled for 2017–2023 from the seven-year country panel (204 countries; GBD 2023). The 2017–2019 baseline was summarised by its mean (μ), sample standard deviation and coefficient of variation (CV), and the natural-variability band was defined around μ with a half-width equal to the largest absolute within-baseline deviation (max |value/μ − 1| over 2017–2019). A pandemic-window year (2020–2023) was classed as outside the band when its deviation from μ exceeded the band half-width; an auxiliary rule flagged declines larger than twice the largest baseline single-year decline. Because the baseline has only three points and GBD point estimates are model-smoothed (baseline CVs are 0.3%–3% for most aetiologies), z-scores were treated as rough references only and the band rule, which does not rely on the standard deviation, was used for classification. Country-level exceedance for Mycoplasma was assessed in the 107 countries with a baseline mean of ≥5 deaths.

Findings. All 26 LRI aetiologies exceeded their 2017–2019 natural-variability bands in both 2020 and 2021, without exception; 25 of 26 remained outside in 2022 (only Legionella spp returned within band) and 23 of 26 were still outside in 2023 (Supplementary Figure S1). The pandemic-window collapse therefore cannot be attributed to natural fluctuation for any node of the LRI spectrum. Depth, however, stratified widely (Supplementary Table S11): respiratory syncytial virus and influenza deviated −65.2% and −62.1% from the 2017–2019 mean in 2021, Streptococcus pneumoniae −33.6% and Mycoplasma −18.9% (7.5 times its largest baseline annual decline of −2.5%; baseline CV 2.0%), whereas tuberculosis—outside the LRI aetiology panel and lacking a three-point baseline—declined only −4.4% from 2019 to 2021. On the V-shape ranking that combines trough depth with the subsequent rebound (Supplementary Table S12), Mycoplasma placed 19th of 26 aetiologies (V-score 33.2), a mid-depth phenotype. Annual deaths of every aetiology moved in near-perfect inverse lockstep with COVID-19 deaths over 2020–2023 (Supplementary Table S13; Spearman ρ = −1.0 for 23 of 26 aetiologies; Mycoplasma Pearson r = −0.901), and at country level 95 of 107 informative countries (88.8%) breached their own Mycoplasma variability bands in 2021, the year of the global COVID-19 mortality peak.

**Supplementary Table S11. Pandemic-window depth gradient against 2017–2019 natural variability: representative causes, global, ages 0–19 years**

| Cause | Reference baseline | 2021 deviation (%) | 2023 deviation (%) |
|---|---|---|---|
| Respiratory syncytial virus | 2017–2019 mean | −65.2 | −48.5 |

| | | | |
|---|---|---|---|
| Influenza | 2017–2019 mean | −62.1 | −45.4 |
| Streptococcus pneumoniae | 2017–2019 mean | −33.6 | −22.4 |
| **Mycoplasma** | **2017–2019 mean** | **−18.9** | **−5.8** |
| Tuberculosis* | 2019 (two-point comparison) | −4.4 | −12.0 |

Note: Values for the four LRI aetiologies are deviations from the 2017–2019 baseline mean (global_natural_variability_metrics.csv, supplementary CSV files). * Tuberculosis is not part of the 26-aetiology LRI panel and lacks a three-point 2017–2019 baseline in the panel (available years 1990, 2010, 2019, 2021, 2023); its entries are simple 2019 to 2021 and 2019 to 2023 percentage changes, shown for reference.

### Supplementary Table S12. V-shape ranking of the 26 LRI aetiologies (top ten plus Mycoplasma), global, ages 0–19 years, 2019–2023

| Rank | Aetiology | Trough year | Trough depth (% vs 2019) | Rebound to 2023 (%) | 2023 vs 2019 (%) | V-score |
|---|---|---|---|---|---|---|
| 1 | Respiratory syncytial virus | 2021 | −65.2 | +48.0 | −48.5 | 113.2 |
| 2 | Influenza | 2021 | −62.6 | +44.4 | −46.1 | 107.0 |
| 3 | Streptococcus pneumoniae | 2021 | −27.1 | +16.7 | −14.9 | 43.8 |
| 4 | Acinetobacter baumannii | 2021 | −23.8 | +17.5 | −10.4 | 41.3 |
| 5 | Haemophilus influenzae | 2021 | −19.9 | +18.3 | −5.2 | 38.2 |
| 6 | Klebsiella pneumoniae | 2021 | −19.8 | +18.4 | −5.0 | 38.2 |
| 7 | Chlamydia spp | 2021 | −19.1 | +18.4 | −4.2 | 37.5 |
| 8 | Enterobacter spp | 2021 | −17.6 | +19.2 | −1.8 | 36.8 |
| 9 | Other bacterial and viral pathogens | 2021 | −17.2 | +19.0 | −1.5 | 36.2 |
| 10 | Escherichia | 2021 | −17.3 | +18.6 | −1.9 | 35.9 |

| | | | | | | |
|---|---|---|---|---|---|---|
| | coli | | | | | |
| **19** | **Mycoplasma** | **2021** | **−17.1** | **+16.1** | **−3.7** | **33.2** |

Note: Trough depth is the percentage change from 2019 to the trough year; rebound is the percentage recovery from the trough to 2023; V-score = |trough depth| + rebound. The Mycoplasma row (rank 19) is bold. The full 26-row ranking is provided as table_global_vshape_ranking.csv (supplementary CSV files).

**Supplementary Table S13. Synchronicity of LRI aetiology deaths with COVID-19 deaths, global, ages 0–19 years, 2020–2023**

| Aetiology | Pearson r vs COVID-19 | Spearman ρ vs COVID-19 |
|---|---|---|
| Influenza | −0.970 | −1.0 |
| Respiratory syncytial virus | −0.960 | −1.0 |
| Other bacterial and viral pathogens | −0.927 | −1.0 |
| Enterobacter spp | −0.927 | −1.0 |
| Group B streptococcus | −0.923 | −1.0 |
| Escherichia coli | −0.923 | −1.0 |
| Haemophilus influenzae | −0.918 | −1.0 |
| Serratia spp. | −0.918 | −1.0 |
| Other Mycobacterium species (non-TB, non-Leprosy) | −0.918 | −1.0 |
| Klebsiella pneumoniae | −0.917 | −1.0 |
| Pseudomonas aeruginosa | −0.917 | −1.0 |
| Other Klebsiella species | −0.915 | −1.0 |
| Chlamydia spp | −0.915 | −1.0 |
| Morganella spp. | −0.913 | −1.0 |
| Group A Streptococcus | −0.913 | −1.0 |
| Citrobacter spp. | −0.912 | −1.0 |
| Aspergillus spp. | −0.910 | −1.0 |
| Staphylococcus aureus | −0.910 | −1.0 |
| Other Acinetobacter species | −0.902 | −1.0 |
| Mycoplasma | −0.901 | −1.0 |

| | | |
|---|---|---|
| Other fungi | −0.898 | −1.0 |
| Other gram-negative bacteria | −0.895 | −1.0 |
| Proteus spp. | −0.890 | −1.0 |
| Acinetobacter baumannii | −0.874 | −0.8 |
| Streptococcus pneumoniae | −0.806 | −0.8 |
| Legionella spp | −0.785 | −0.8 |

Note: Pearson and Spearman correlations between each aetiology’s global annual deaths and global COVID-19 annual deaths (ages 0–19 years) over 2020–2023 (four annual points). Values are descriptive; with n = 4 no significance testing was performed. Rows are ordered by the Pearson coefficient (table_covid_synchronicity.csv).

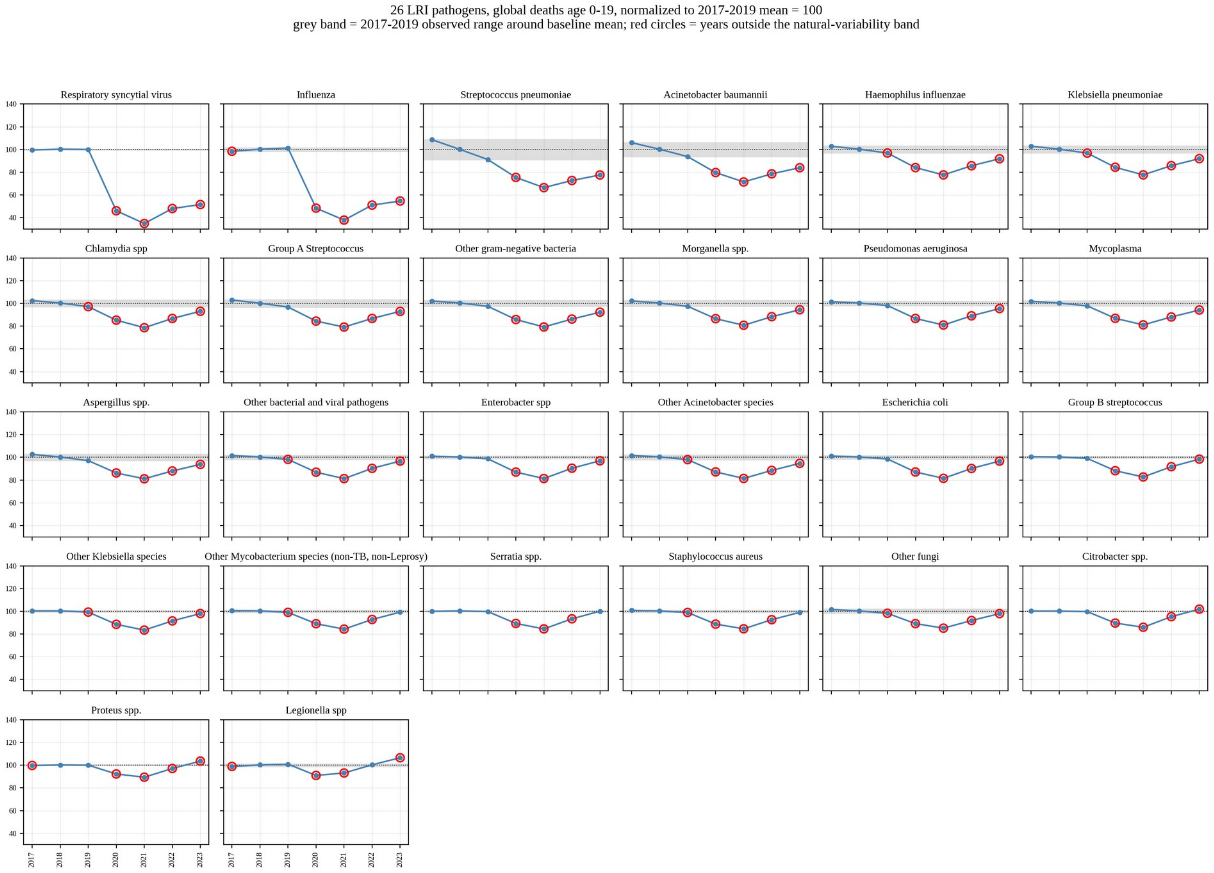


**Supplementary Figure S1.** Natural variability versus pandemic-window deviation for the 26 LRI aetiologies, global, ages 0–19 years, 2017–2023. Annual deaths normalised to the 2017–2019 baseline mean (= 100); the grey band is the observed 2017–2019 variability band around the baseline mean and red circles mark years outside the band. Every aetiology left its band in 2020 and 2021, and most remained outside through 2023. Source data: global_natural_variability_metrics.csv (supplementary CSV files).

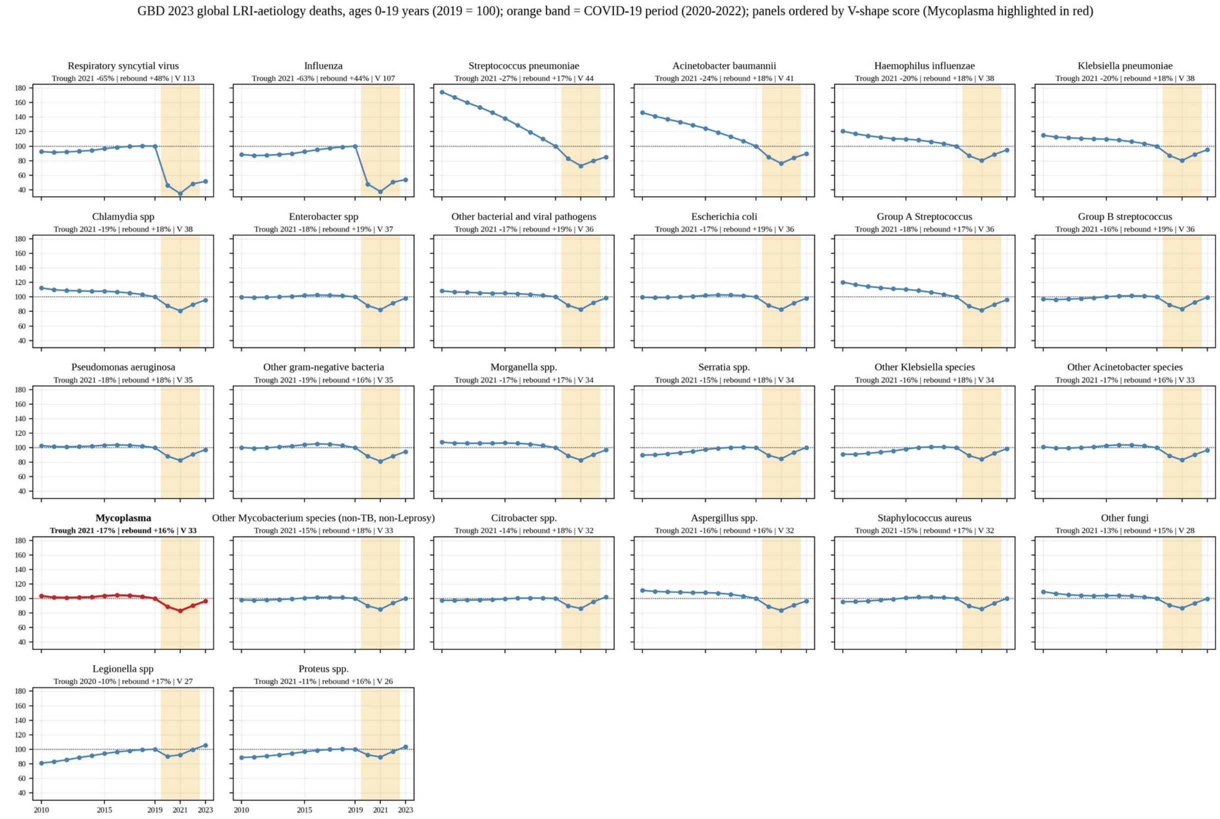


**Supplementary Figure S2.** Pandemic-window V-shape trajectories of the 26 LRI aetiologies, global, ages 0–19 years, 2010–2023 (2019 = 100), ordered by V-shape score (trough depth plus rebound to 2023). The orange band marks 2020–2022 and Mycoplasma is highlighted in red; panel titles give the trough year and depth, the rebound to 2023 and the V-score. Source data: x_global_pathogen_year_0_19_deaths_29.csv and table_global_vshape_ranking.csv (supplementary CSV files).
